# Estimation of hospital catchment populations using data on patient hospital use in France

**DOI:** 10.64898/2026.04.28.26351911

**Authors:** George Shirreff, Cécile Chauvel, Jean-Sébastien Casalegno, Philippe Vanhems, Cédric Dananché, Abdesslam Redjaline, Karim Tazarourte, Marta Nunes

## Abstract

**Background:** Estimates of disease burden from hospital data require well-informed estimates of the size of the catchment population. Data on patient flows from residential areas to a hospital can be used to estimate detailed catchment populations by age, year and type of hospital visit.

**Methods:** Catchment populations were estimated for hospitals throughout France using a proportional flow approach. Data on hospital use and patient residence were accessed from the Agence Technique de l’Information sur l’Hospitalisation (ATIH). For patients coming from each administrative area, we calculated a preference for each hospital, and combined this with population data for the area to estimate the catchment population of each hospital. For one hospital group, we compared this with data on emergency visits, and data from a retrospective cohort study.

**Results:** Estimated catchment population by hospital group ranged from 4 million per year for Assistance Publique – Hôpitaux de Paris (AP-HP) downwards, with the catchment population strongly reflecting geographic proximity and hospital scale. The type of hospital substantially impacted the size of the catchment area. In the analysis of a single hospital group, the size of the catchment population varied widely with the diagnostic categories associated with the hospital visit. Emergency visits represented a smaller catchment population. The estimated proportional contribution of different departments to the estimated catchment population was similar to their contribution to observed hospital admissions. Incidence rates for a respiratory virus using this catchment population estimation method were consistent with national incidence rates.

**Conclusions:** This study demonstrates the consistency of the proportional flow framework when applied to appropriate data on hospital usage. The study provides catchment populations for each hospital in France which can be used for burden estimates such as incidence rates, as well as providing insight into the catchment populations served. Analysis at the department geographic level provided an appropriate balance between detail of analysis and the need to mask data for anonymisation. Further analysis should explore how the size of the catchment area corresponds to the associated travel time to the hospital in question.

## Introduction

Hospital-based data are widely used to estimate disease burden, healthcare utilisation, service demand and costs. Counts of emergency visits and hospitalisations frequently serve as numerators in epidemiological analyses. However, translating hospital case counts into population-level disease incidence requires accurate definition of the population from which these cases arise (1). This denominator is typically conceptualised as the hospital catchment population, who would attend a given hospital if they required care (2). Mis-specification of this denominator can bias incidence estimates and distort comparisons across hospitals, regions or time periods.

Different approaches have been used to estimating hospital catchment populations. Some studies have identified the catchment area based purely on geographic information, either by identifying areas falling within circles of a fixed distance from the hospital (3), or a within a fixed travel time (4). While intuitive, these approaches assume that patients attend their nearest facility and cannot easily account for hospital ranges, specialisation or referral patterns. However, the use of different facilities can be understood using patient flow i.e. data on hospital usage by the population within a geographic area. For example, a catchment area can be defined by identifying the administrative areas which contain above a fixed percentage of patients’ addresses (5,6). This method, referred to as first-past-the-post, assigns the entire population of the chosen administrative areas to the catchment of the most popular hospital (7), but does not reflect the overlapping service areas observed in practice.

Alternatively, the proportional flow model allocates the population residing in a geographic area to be assigned to multiple hospital catchments according to the observed utilisation patterns (8). Further refinements are possible to this approach, such as those which begin by assuming that the geographic area in which the hospital is situated form the hospital catchment areas, and then refine the value for each hospital using in-flows, out-flows and overall admission rates (2). Although this framework has strong conceptual foundations, national-scale applications using comprehensive administrative hospital data remain limited.

In this analysis, we analysed the internal consistency of the proportional flow model across different types of source data and validated the results against a clinical cohort. The objective of this study is to provide a scalable methodological framework to strengthen hospital-based surveillance and burden estimation.

## Methods

To implement the proportional flow method, we used national or regional data on hospital use to estimate for each administrative area the proportion of the population preferring each hospital. This was then combined with population data to estimate the size and distribution of the catchment population for each hospital. We conducted analyses across all hospitals and with a focus on one specific hospital group, the Hospices Civils de Lyon (HCL). We validated the analysis by comparison with a large retrospective cohort of patients within the hospital group.

### Hospitalisation and emergency department data

Data are held by Programme de Médicalisation des Systèmes d’Information (PMSI) on the numbers of hospitalised patients coming to each hospital from each geographic area of residence. These data were acquired via the ATIH using Activité MCO, a query tool which covers hospitalisations under medicine, surgery and obstetrics. We extracted annual counts of patients hospitalised for at least for one night, disaggregated by year of hospitalisation, age group, major diagnostic category (9), and geographic provenance (region, department or PMSI code). For each stratum, counts were obtained both for patients attending any hospital as well as patients attending a particular hospital group, as identified by the Fichier National des Établissements Sanitaires et Sociaux (FINESS) number. Cells within the dataset which contained fewer than 11 patient admissions per year, per hospital and per patient characteristic were automatically anonymised by the query tool which aggregated all such cells into a single row. Hospital categories for each FINESS number were obtained from FINESS (10). Analyses focused in particular on the HCL (FINESS number 690784137), with 12 individual hospitals in the Rhône department, Auvergne-Rhône-Alpes (ARA) region, and one hospital outside the region.

Data on the number of patients with emergency department visits were obtained for the ARA region from Urg’ARA, stratified by year, age group and postcode of residence. Counts of emergency visits to any hospital in the region, and to the HCL group specifically, as well as the proportion visiting the HCL. Cells within this dataset with fewer than 5 patient admissions were masked for anonymisation meaning the actual number visiting HCL was not given, but the proportion of all emergency visits which went to HCL was still provided if available.

### Geographic levels

Geographic provenance of patients was defined at three levels: region, department, and PMSI areas defined by a PMSI code. PMSI codes correspond to postcodes when the area defined by the postcode exceeds 1000 residents, or to aggregations of two or more postcodes (11). In our principal analysis, we restricted the population to Metropolitan France (mainland France and nearby islands including Corsica). We also conducted similar analyses of overseas regions, which included Guadeloupe, Martinique, French Guyana, La Reunion and Mayotte. Geographic boundary shapefiles for regions, departments and postcodes, including their area in km^2^, were accessed from publicly available records (12–14). The value for the entire surface area of Metropolitan France was obtained from INSEE, adding together the area excluding Paris (and the area of Paris separately (15,16).

### Population data

Annual population sizes by age were obtained from the Institut National de la Statistique et des Etudes Economiques (INSEE). Population estimates for regions and departments were accessed as annual counts in 5-year age bands (17,18). Where finer age stratification was required, proportions of 1-year age bands within 5-year bands were derived from national 1-year age distributions (19) and applied to regional and departmental totals. Population data by age and INSEE geographic code for 2021 was accessed from INSEE (20) and linked to postcodes using official correspondence tables (21). PMSI codes were linked to postcode-level population data using correspondence tables published by Agence Technique de l’Information sur l’Hospitalisation (ATIH) (22).

### Calculation of population in catchment area

We estimated the catchment populations and catchment area using a simple proportional flow approach described by Bailey (8) but extended to include attributes of the population beyond its geographic area allowing stratification by age, year and visit characteristics.

For each group of patients *M* with a set of characteristics (geographic provenance *g*, year of hospitalisation *y*, age group *a*, and other characteristics θ including major diagnostic category or emergency visit), the preference of those patients for Hospital A is the proportion of patients attending Hospital F as a proportion of all patients attending any hospital. The contribution C of each geographic area to the catchment area of Hospital F was then calculated by multiplying the preference by the population size *N*:

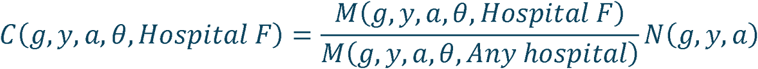

A similar approach was taken to estimate the catchment area of a given hospital. The contribution A of each geographic area to the total catchment area was calculated by multiplying the patient preference by the surface area *S* of the geographic area in km^2^.

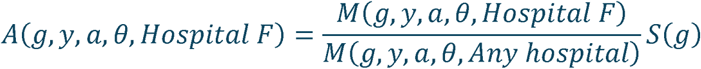

Summing contributions of *C,* and of *A,* across all geographic areas yielded respectively the overall catchment population, and catchment area, of each hospital according to a characteristic of interest. Analyses were conducted separately at the region, department and PMSI code levels to assess the impact of geographic resolution on denominator estimates. Anonymised cells in the dataset were excluded from the analysis.

We also estimated the relative contribution of each geographic area *g* to the catchment population by dividing the estimate by the sum of all catchment contributions across all geographic areas, as follows:

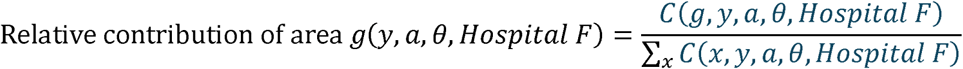

For displaying data on maps, the catchment density was calculated by dividing the population by the area in km^2^.

### Access to local services

We measured the level of access to local health services by calculating the proportion of the population in a given department who fall within the catchment area of a hospital in another department. The department of the hospital was derived from the first two characters of the FINESS code. The proportions were calculated per year and then averaged over all years available.

### Respiratory cohort from the HCL

Validation of the proportional flow estimates was conducted using a retrospective cohort at the HCL. Patients were included if they visited the emergency department or were hospitalised at one of the hospitals in the HCL group between 1 January 2017 and 31 December 2024 with an acute respiratory infection. This was defined by meeting at least one of the following criteria: a positive PCR test for a respiratory virus (influenza, respiratory syncytial virus, SARS-CoV-2, enterovirus/rhinovirus, human metapneumovirus, adenovirus, parainfluenza viruses or non-SARS coronaviruses); or an International Classification of Diseases version 10 (ICD-10) diagnostic code consistent with acute respiratory infection (list of codes in Supplementary Information). For each patient, age, date of admission and department of residence were extracted. Visits were classified as emergency department only or hospitalisation.

For both hospitalisation and emergency visit and for each year and age group separately, the proportion of patients coming from each geographic department was calculated as the number of patients from that department over the total number admitted. These proportions were used to compare the geographic distribution of observed respiratory admissions with the estimated catchment population distribution derived from the proportional flow model.

In order to validate the denominators as measures of disease incidence, we calculated the incidence per 1000 population of respiratory syncytial virus (RSV) in the respiratory cohort. In order to compare directly with a national analysis in children <1 year of age (23), we defined an infant <1 year with RSV as one assigned an explicit RSV ICD-10 code (J12.1, J20.5 or J21.0) at any time of year, or a non-specific ICD-10 code (B97.4, J20.8, J20.9, J21.8, J21.9, J45) during the season from October through March. The incidence per 1000 population was calculated by dividing the number of cases in each year with the yearly catchment population for children <1 year.

## Results

### Analysis across all hospitals

Between 2016 and 2025, annual overnight hospitalisations recorded in the national administrative dataset ranged from 8.7 to 10.5 million (Table 1). Across all years, approximately 96% of patients had a recorded residence within Metropolitan France, 3% resided in overseas regions, and 0.5% had missing residence information.

**Table 1.**
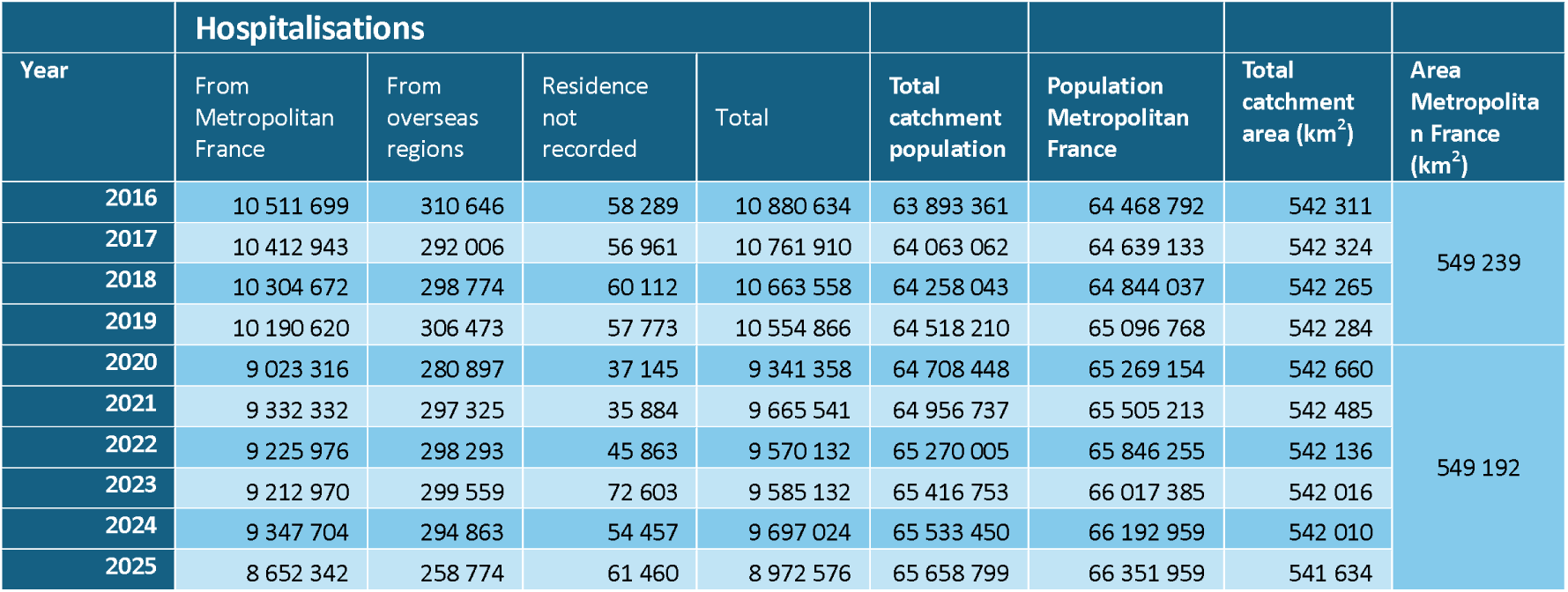
Total overnight hospitalisations recorded in PMSI by year. The catchment population is the total across all estimates calculated at the resolution of the department of residence and the hospital group FINESS number.

Application of the proportional flow model generated catchment population estimates for all hospital groups in Metropolitan France. Catchment size varied substantially according to hospital scale. Among the largest ten hospital groups, which were all major university hospital centres, estimated annual catchment populations ranged downwards from approximately 4.0 million for Assistance Publique–Hôpitaux de Paris (AP-HP) which covered an estimated catchment area of 5000 km^2^ (Table 2). For the HCL, the estimated catchment population was approximately 1.1 million residents annually, but the similar catchment area (4400 km^2^) implies a catchment area of lower density. The largest catchment area was for CHU Toulouse. We also provide the catchments of the largest 200 hospital groups serving Metropolitan France (Supplementary Table 1), the largest 100 individual hospitals serving Metropolitan France (Supplementary Table 2) and the largest 50 hospital groups serving overseas regions (Supplementary Table 3).

**Table 2.** Average annual number of hospitalisations and size of estimated catchment population for the ten largest hospital groups, by number of hospitalisations, over the period 2016-2025. CHU=University hospital centre.

| FINESS – Hospital group | Annual hospitalisations | Estimated catchment population | Estimated catchment area (km<sup>2</sup>) |
| --- | --- | --- | --- |
| 750712184 - Assistance Publique-Hôpitaux de Paris (AP-HP) | 525 515 | 3 999 251 | 4 999 |
| 690781810 – Hospices Civils de Lyon (HCL) | 154 342 | 1 097 187 | 4 407 |
| 130786049 - Assistance Publique-Hôpitaux de Marseille (AP-HM) | 112 620 | 709 650 | 3 703 |
| 310781406 - CHU Toulouse | 112 118 | 799 420 | 7 409 |
| 330781196 - CHU de Bordeaux | 100 988 | 704 499 | 6 036 |
| 590780193 - Centre hospitalier régional universitaire Lille | 92 191 | 567 661 | 1 614 |
| 670780055 - CHU de Strasbourg | 84 053 | 577 094 | 2 592 |
| 440000289 - CHU de Nantes | 74 331 | 588 517 | 3 320 |
| 340780477 - CHU Montpellier | 71 919 | 489 841 | 3 946 |
| 540023264 - CHU de Nancy | 69 514 | 444 182 | 4 582 |

The high proportion of admissions with known geographic provenance allowed estimation of hospital preference proportions for nearly the entire population of Metropolitan France. The total estimated catchment population across all hospital groups increased gradually over time and closely matched annual population estimates for Metropolitan France (Table 1).

Geographic distribution of catchment contributions reflected both proximity and hospital scale. While most hospital groups drew the majority of their catchment population from surrounding departments, larger centres generally had a wider reach (Figure 1), illustrating the overlapping and non-exclusive nature of hospital service areas.

**Figure 1.**
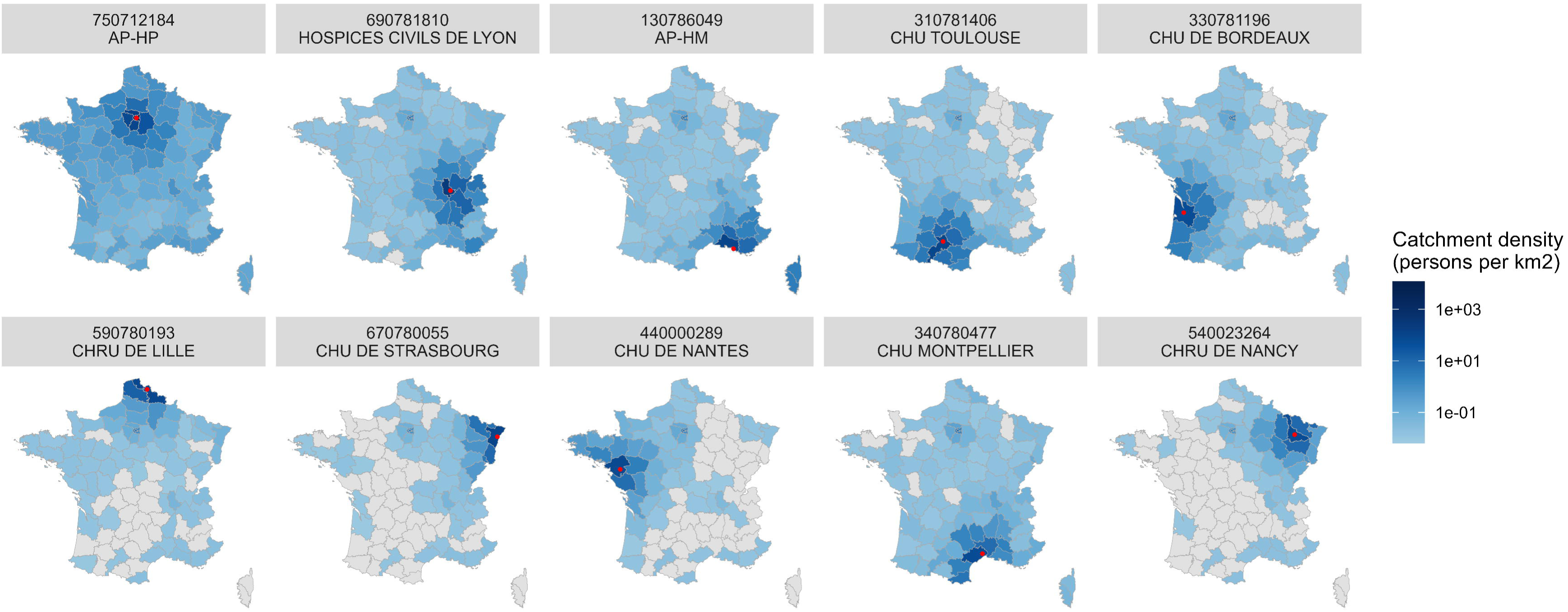
Estimated density of the catchment population coming from each department to the ten largest hospitals. Hospital group FINESS number and name above each panel. Departments in grey contributed fewer than 10 hospitalisation visits to that hospital in any of the years considered. The red point represents the main address of the hospital group.

All of the hospitals represented in Table 2 are categorised as regional hospital centres. We examined the distribution of catchment populations and catchment areas by this and other categories of hospitals (Figure 2). This demonstrates not only the differences in magnitude between hospital categories, with regional hospital centres (CHR) having the largest catchment populations, but also differences in variance, with hospital centres (CH) having a wider range of catchment areas and populations.

**Figure 2.**
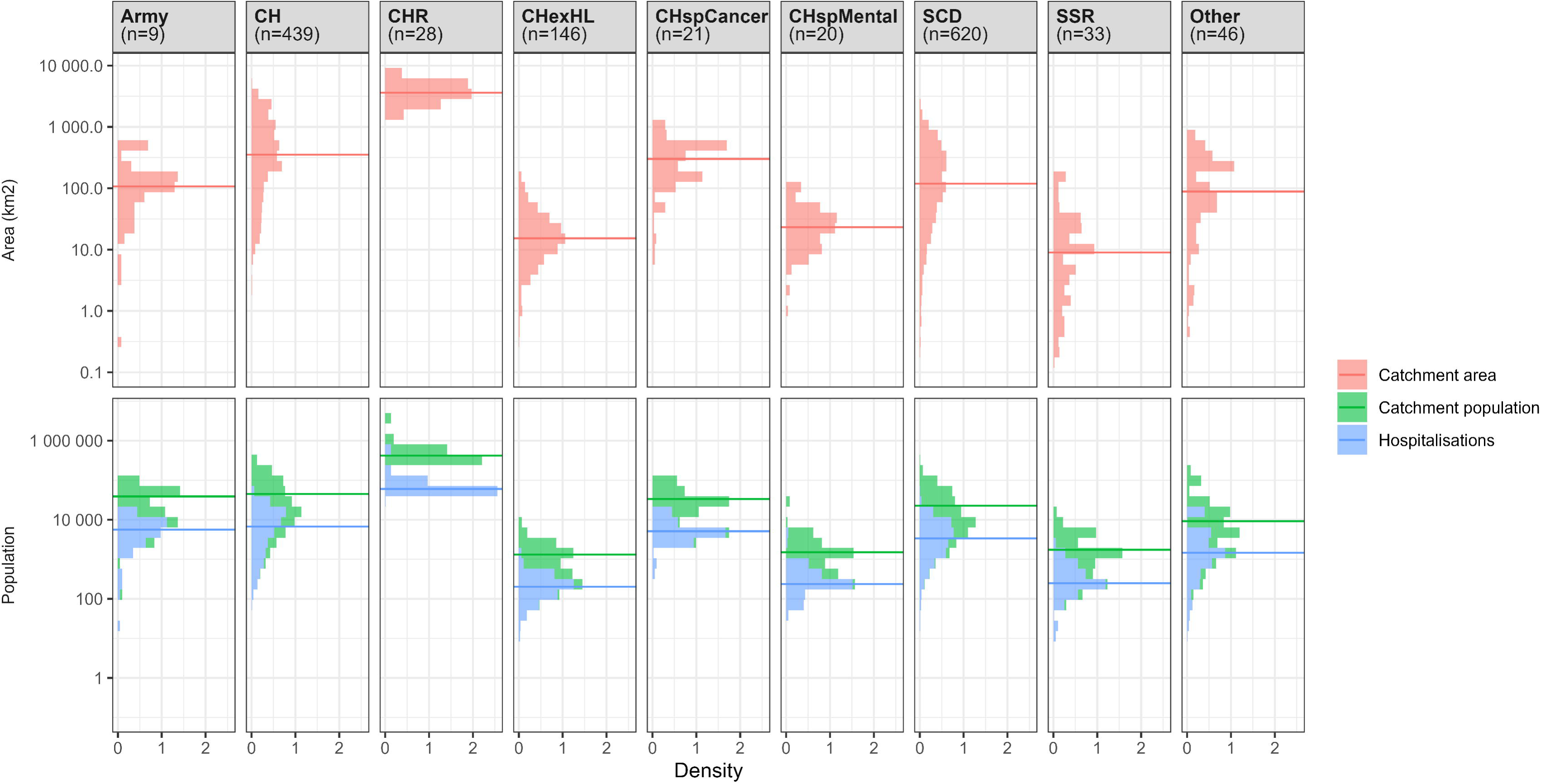
The distribution of catchment areas (top row) and catchment populations (bottom row) according to the type of hospital (columns). The distribution of the number of hospitalisations is also included in the bottom row. The horizontal lines indicate the median for that hospital category. CH=Hospital centre, CHexHL=Hospital centre, formally a local hospital, CHR=Regional hospital centre, CHspCancer=Hospital centre specialising in cancer, CHspMental=Hospital centre specialising in mental health, SCD=Acute care hospital, SSR=Rehabilitation and recuperative care.

In order to measure how access to healthcare varied geographically, we examined the proportion of residents of each department who sought care in another department, for various different diagnostic categories (Figure 3). This demonstrates that for mental health services patients were largely hospitalised within their own department, whereas for organ transplants patients tended to leave the department. This also indicates areas where specialist services such as burns and organ transplants are not available locally.

**Figure 3.**
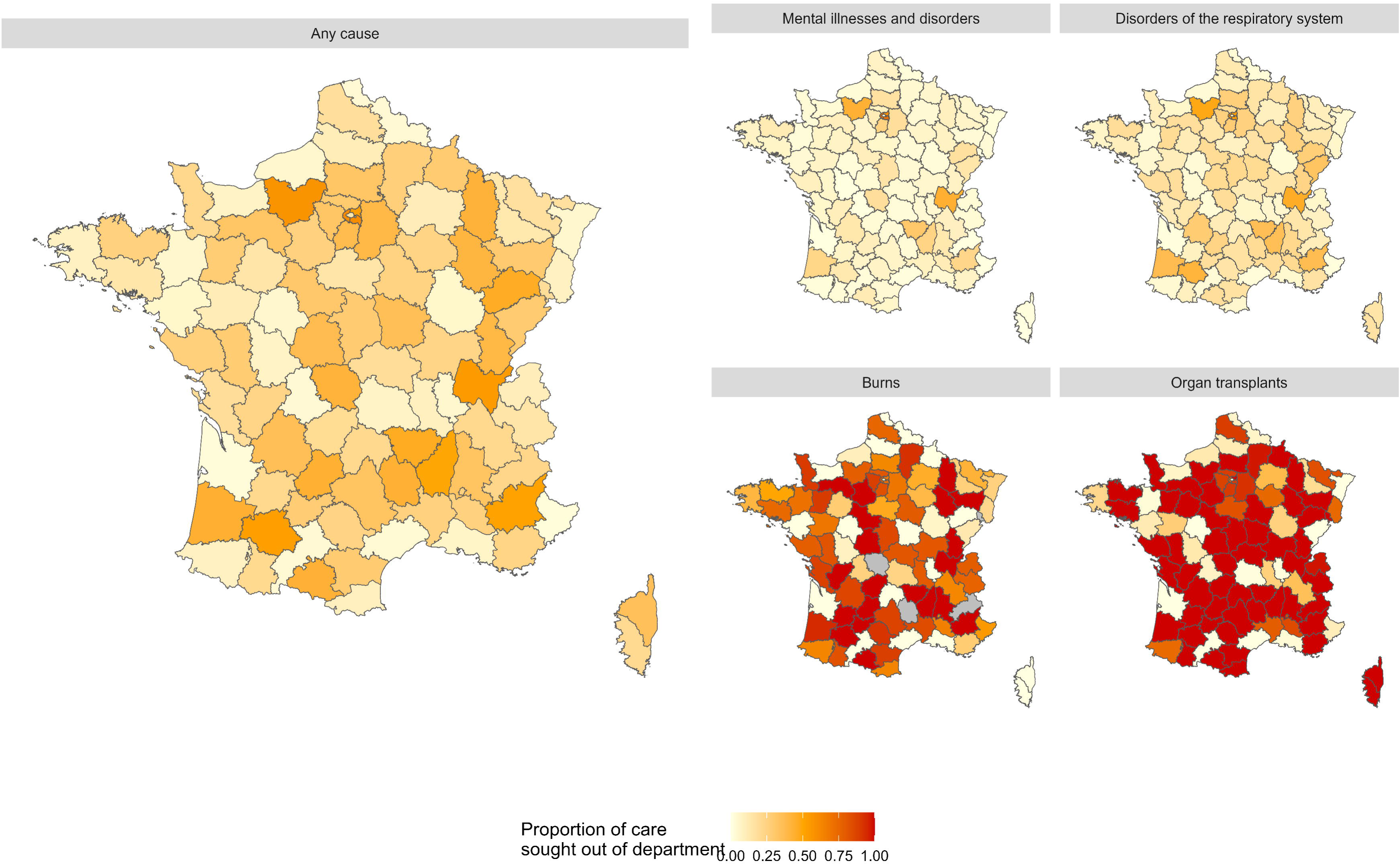
Local health access in each geographic department, defined as the proportion of the resident population who sought care in another department. This is shown for any reason for hospitalisation on the left, and by a representative selection of specific reasons for hospitalisations on the four panels on the right. The grey areas were missing values, where there were insufficient data to calculate a proportion.

### Detailed case study: HCL

To explore how the assumptions made in the analysis affected estimates of catchment population, we conducted detailed analysis of a specific hospital group.

We evaluated the influence of geographic aggregation on estimated catchment populations by comparing total catchment populations as estimated at different geographic levels of resolution. Catchment population estimates varied according to resolution (Figure 4). Analyses conducted at the PMSI code-level yielded smaller overall catchment populations than for those at the region or department level, for each age group separately and all groups combined. This was likely attributable to more frequent aggregation due to anonymisation rules in the national administrative dataset. There were smaller differences in catchment populations when conducted at the regional or departmental level, with the regional values being smaller since 2021 in younger age groups. Figure 4 also demonstrates that the size of the catchment population did not change substantially over the course of the COVID-19 pandemic, suggesting that although overall numbers of hospitalisations nationally decreased sharply in 2020 (Table 1), this trend did not result in a change in patient preference for the HCL during this time.

**Figure 4.**
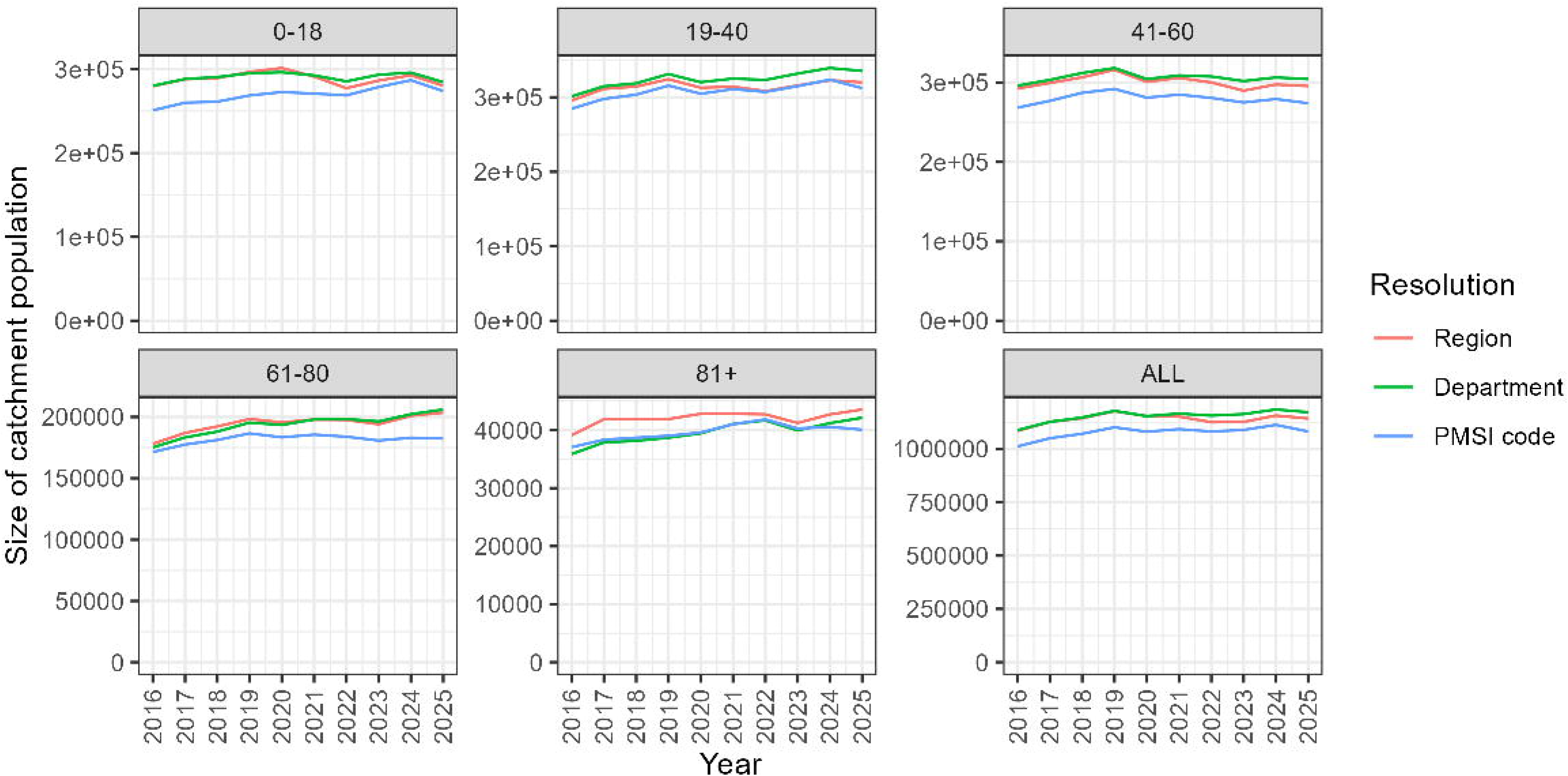
Estimated annual catchment population sizes by year and age group in the HCL. Analysis was conducted either at the resolution of the region, department or PMSI code.

Higher numbers of PMSI codes were excluded from the analysis due to sending fewer than 11 patients to the HCL per year and therefore being excluded from the dataset at this level of aggregation. The proportion of admissions affected by masking increased as geographic resolution become finer. At the regional level with age stratification, 0.00% and 0.04% of overall admissions and HCL admissions were masked, respectively; at the department level the respective proportions of masked admissions were 0.00% and 0.59%; and at the PMSI code level, 0.23% and 6.65% were masked. As a result, finer geographic resolution increased spatial specificity but reduced completeness of denominator estimation due to masked low-volume flows. In contrast, coarser aggregation reduced masking but potentially obscured heterogeneity in hospital preference patterns.

To further illustrate the geographic distribution of the estimated catchment population, Figure 5 presents the density of the HCL catchment in 2024 across all ages, displayed separately at the regional, departmental and PMSI code levels. The catchment area extended across Metropolitan France, with all regions and most departments contributing measurable population shares to the HCL. However, at the PMSI level, numerous small geographic areas either did not contribute or were excluded due to the anonymisation threshold, including even some in relatively close proximity to the HCL. Although the catchment population was drawn from a wide geographical area, its total remained considerably smaller than either the resident population of the Rhône department or the Lyon Metropolitan area in which the HCL is located (respectively 1 907 982 and 1 433 613 in 2022 (24,25)), reflecting the fact that only a proportion of the residents of these areas utilise the HCL for inpatient care.

**Figure 5.**
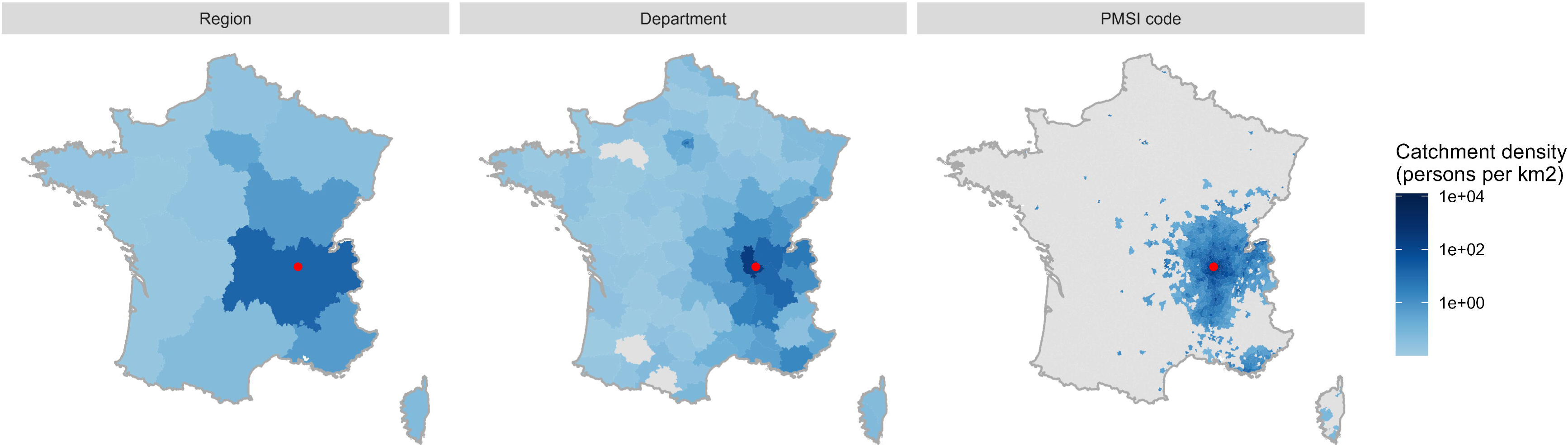
Density of the HCL catchment population in 2024 across all ages, by region, department and PMSI code. The colour code is shown on a log scale. Grey areas are those where the number of cases going to HCL that year are lower than the anonymity threshold.

We also examined the catchment area of the different sites of the HCL, which are shown in Figure 6, which demonstrates the overlap in catchment area between the different sites. Distinct spatial patterns were observed across the sites, demonstrating the wide reach of the largest hospitals, the relatively local reach of the smaller sites, and in one instance the wide but rarified area served by a specialist paediatric cancer hospital (IHOP).

**Figure 6.**
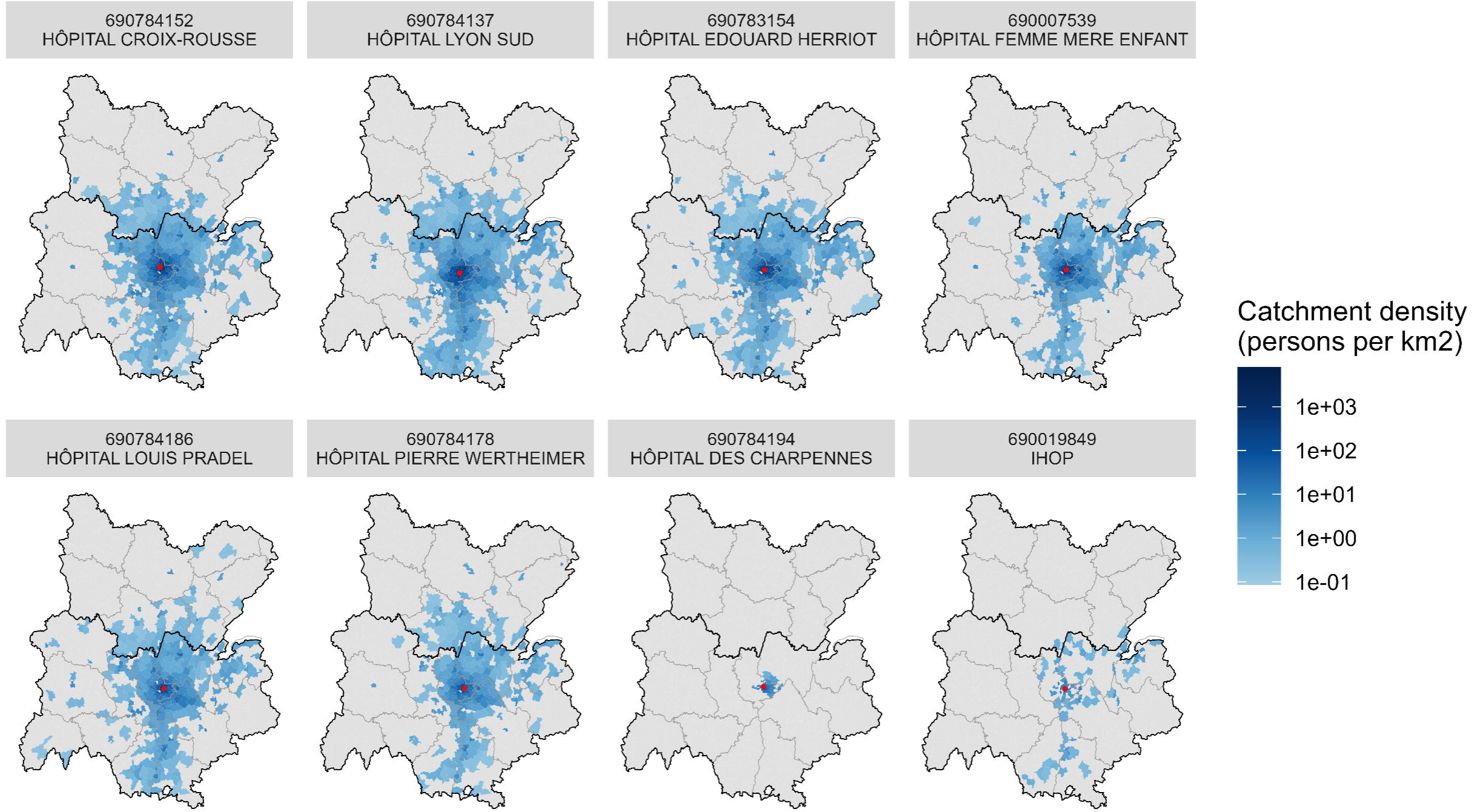
Estimated average annual catchment population by PMSI code for the 8 largest Lyon-based sites of the HCL group in the two regions accounting for most of the catchment area, these being ARA (lower) and Bourgogne-Franche-Comté (upper). Black lines delineate the two regions, and lighter grey lines indicate departmental boundaries. The red point is the location of the site in question.

We obtained age-specific catchment population estimates for hospitalisation at the HCL between 2016 and 2025 (Table 3). Age stratification meaningfully influenced denominator estimates. When analyses were conducted without age disaggregation, the estimated overall catchment population for the HCL (1.1 million, Table 2) was lower than the sum of the catchment populations calculated separately for each age group (1.24 million, Table 3).

**Table 3.**
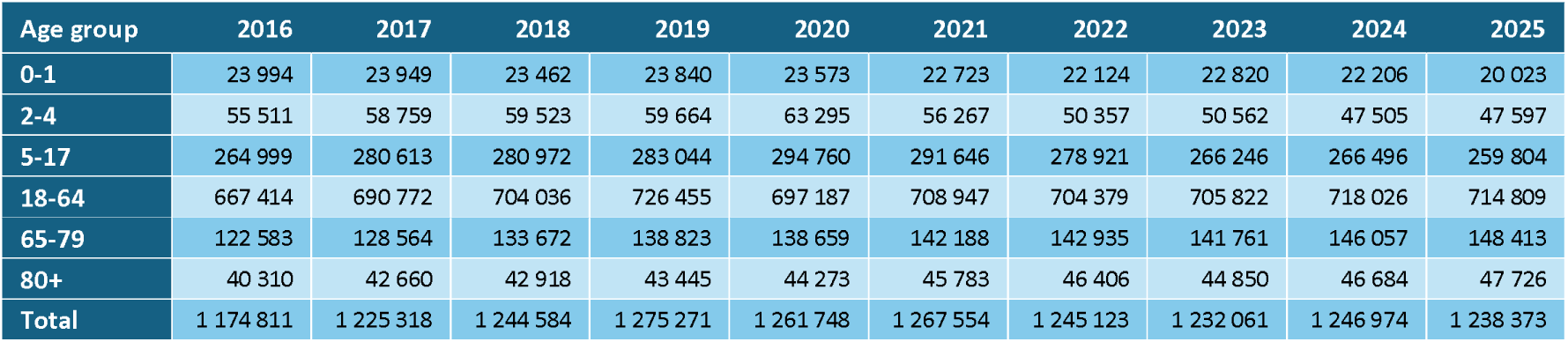
Estimated HCL catchment population sizes by year and age group. Analysis was conducted on data with department level resolution.

| Age group | 2016 | 2017 | 2018 | 2019 | 2020 | 2021 | 2022 | 2023 | 2024 | 2025 |
| --- | --- | --- | --- | --- | --- | --- | --- | --- | --- | --- |
| 0-1 | 23 994 | 23 949 | 23 462 | 23 840 | 23 573 | 22 723 | 22 124 | 22 820 | 22 206 | 20 023 |
| 2-4 | 55 511 | 58 759 | 59 523 | 59 664 | 63 295 | 56 267 | 50 357 | 50 562 | 47 505 | 47 597 |
| 5-17 | 264 999 | 280 613 | 280 972 | 283 044 | 294 760 | 291 646 | 278 921 | 266 246 | 266 496 | 259 804 |
| 18-64 | 667 414 | 690 772 | 704 036 | 726 455 | 697 187 | 708 947 | 704 379 | 705 822 | 718 026 | 714 809 |
| 65-79 | 122 583 | 128 564 | 133 672 | 138 823 | 138 659 | 142 188 | 142 935 | 141 761 | 146 057 | 148 413 |
| 80+ | 40 310 | 42 660 | 42 918 | 43 445 | 44 273 | 45 783 | 46 406 | 44 850 | 46 684 | 47 726 |
| Total | 1 174 811 | 1 225 318 | 1 244 584 | 1 275 271 | 1 261 748 | 1 267 554 | 1 245 123 | 1 232 061 | 1 246 974 | 1 238 373 |

### Analysis by causes of admission

Catchment population size varied substantially according to the clinical characteristics of hospitalisation visits. Using department-level geographic resolution, we estimated yearly average HCL catchment populations by major diagnostic categories (Table 4). While the estimated catchment populations for some conditions such as burns and organ transplants were substantially higher than the equivalent for all causes, many such as mental disorders related to drug use and diseases of the male genital system had a much smaller catchment population.

**Table 4.** Average annual catchment population of the HCL across all ages by diagnostic category.

| Diagnostic category | Catchment population | Catchment area (km<sup>2</sup>) |
| --- | --- | --- |
| Burns | 4 440 889 | 31 129 |
| Organ transplants | 3 571 779 | 21 771 |
| Severe multiple trauma | 2 022 305 | 8 359 |
| Myeloproliferative disorders and neoplasms of uncertain or diffuse site | 1 764 839 | 9 103 |
| Disorders of the eye | 1 553 098 | 3 032 |
| Diseases due to HIV infection | 1 510 090 | 6 413 |
| Disorders of the nervous system | 1 422 773 | 6 320 |
| Factors influencing health status and other reasons for use of health services | 1 336 058 | 6 109 |
| Disorders of the ears, nose, throat, mouth and teeth | 1 288 286 | 4 366 |
| Disorders of the circulatory system | 1 188 730 | 5 062 |
| Injuries, allergies and poisonings | 1 162 589 | 4 020 |
| Disorders of the blood and hematopoietic organs | 1 122 741 | 4 590 |
| Endocrine, metabolic and nutritional disorders | 1 098 901 | 3 058 |
| Disorders of the digestive tract | 1 097 913 | 4 431 |
| Infectious and parasitic diseases | 1 092 854 | 3 856 |
| Disorders of the respiratory system | 1 040 167 | 3 504 |
| Disorders of the hepatobiliary system and pancreas | 1 037 492 | 4 475 |
| Pathological pregnancies, childbirth and postpartum disorders | 1 004 673 | 4 121 |
| Mental illnesses and disorders | 946 836 | 3 737 |
| Newborns, premature infants and conditions of the perinatal period | 934 156 | 3 547 |
| Disorders and injuries of the musculoskeletal system and connective tissue | 918 397 | 2 619 |
| Disorders of the kidney and urinary tract | 902 613 | 2 660 |
| Disorders of the female genital system | 891 499 | 3 237 |
| Disorders of the skin, subcutaneous tissue and breasts | 789 172 | 3 212 |
| Disorders of the male genital system | 718 901 | 1 565 |
| Organic mental disorders related to or induced by drug use | 660 142 | 2 778 |

We estimated the size of catchment populations for emergency department visits using a regional data source. Patient preference was estimated from the ratio of emergency department visits at HCL to emergency department visits at any hospital in ARA. The estimated catchment population by age group and year are shown in Table 5. Emergency visit-based catchment populations were consistently smaller than those derived from hospitalisations, suggesting more geographically localised utilisation patterns for emergency care.

**Table 5.** Catchment population by age group and year for emergency department visits to HCL from patients with provenance in the ARA region. For comparison we include the equivalent totals for hospitalisation visits with provenance in ARA.

| Age group | Visit | Region | 2019 | 2020 | 2021 | 2022 | 2023 | 2024 |
| --- | --- | --- | --- | --- | --- | --- | --- | --- |
| 0-1 | Emergency | ARA | 32 637 | 34 779 | 33 853 | 32 575 | 32 498 | 29 826 |
| 2-4 | Emergency | ARA | 48 292 | 49 160 | 50 360 | 48 049 | 45 556 | 45 850 |
| 5-17 | Emergency | ARA | 154 626 | 166 311 | 169 319 | 172 366 | 168 099 | 171 153 |
| 18-64 | Emergency | ARA | 361 611 | 355 758 | 374 311 | 386 617 | 394 222 | 384 831 |
| 65-79 | Emergency | ARA | 74 575 | 77 226 | 79 379 | 84 776 | 84 352 | 81 679 |
| 80+ | Emergency | ARA | 38 320 | 42 066 | 42 722 | 44 176 | 44 880 | 43 612 |
| All | Emergency | ARA | 710 061 | 725 301 | 749 943 | 768 558 | 769 606 | 756 952 |
| All | Hospitalisation | ARA | 1 139 789 | 1 131 819 | 1 138 971 | 1 125 753 | 1 119 016 | 1 131 261 |

### Validation using a respiratory cohort from the HCL

To evaluate the validity of the proportional flow-derived denominators for infectious disease surveillance, we compared estimated catchment population distributions, estimated from all causes of hospitalisation, with observed hospital admissions and emergency visits from a respiratory infection cohort at the HCL. In total, 99 680 patients admitted to hospital and 40 409 visiting emergency between 2017 and 2024 inclusive were identified in the respiratory cohort and contributed to this validation analysis.

For each combination of department of residence, age group and year, we compared the proportion of the estimated catchment population residing in that department with the proportion of respiratory admissions originating from the same department (Figure 7). Across strata, these proportions were strongly correlated, with a Pearson correlation coefficient of 0.91 (p<0.001) for hospital admissions and 0.89 (p<0.001) for emergency visits. The proportions close to 1 on both graphs represent individuals coming from the Rhône department in which the HCL is situated. The close agreement between denominator-derived geographic distributions and independently observed respiratory admissions supports the validity of the proportional flow framework for hospital-based infectious disease surveillance. Importantly, this agreement persisted across multiple age groups and epidemic seasons characterised by varying patterns of respiratory virus circulation and healthcare utilisation.

**Figure 7.**
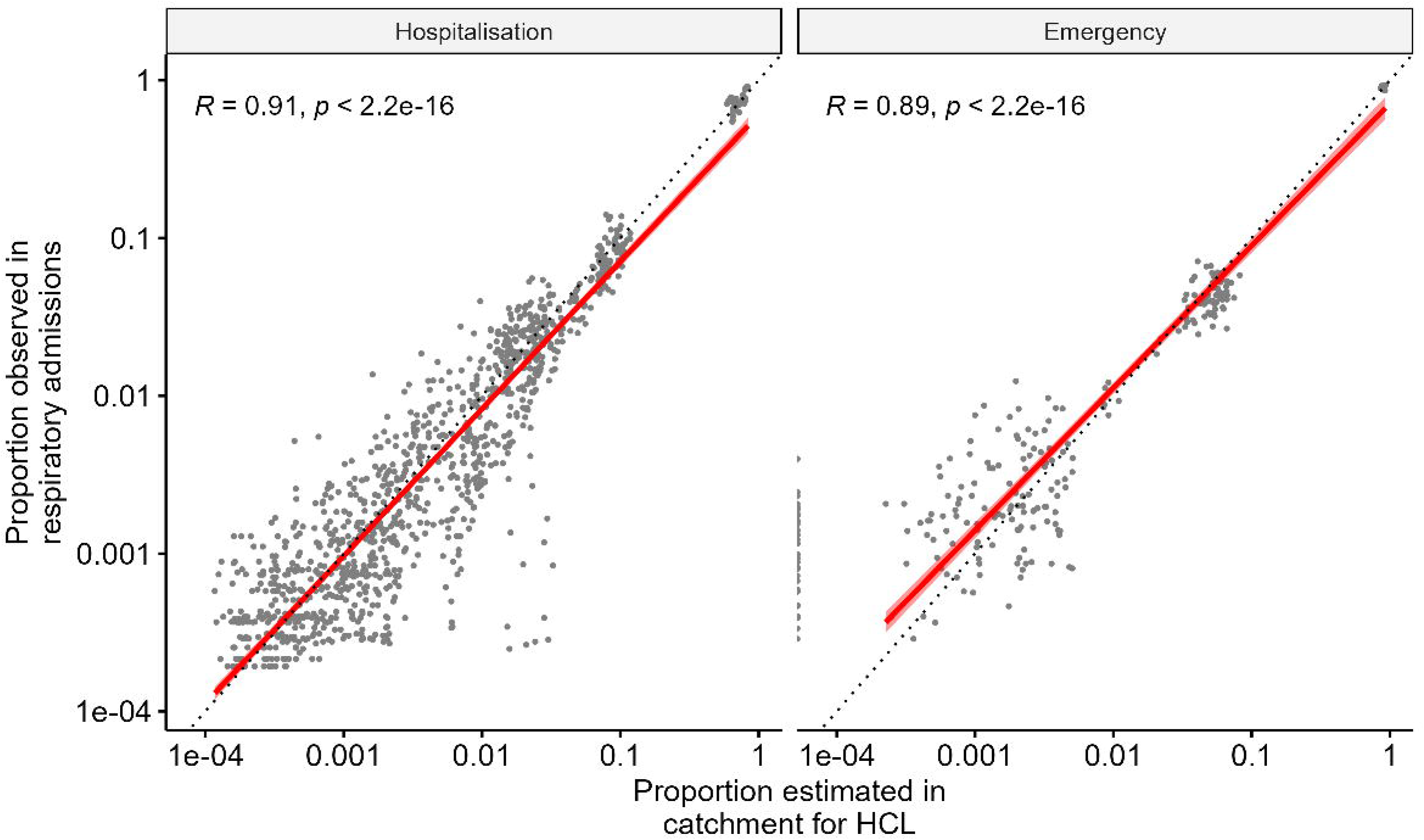
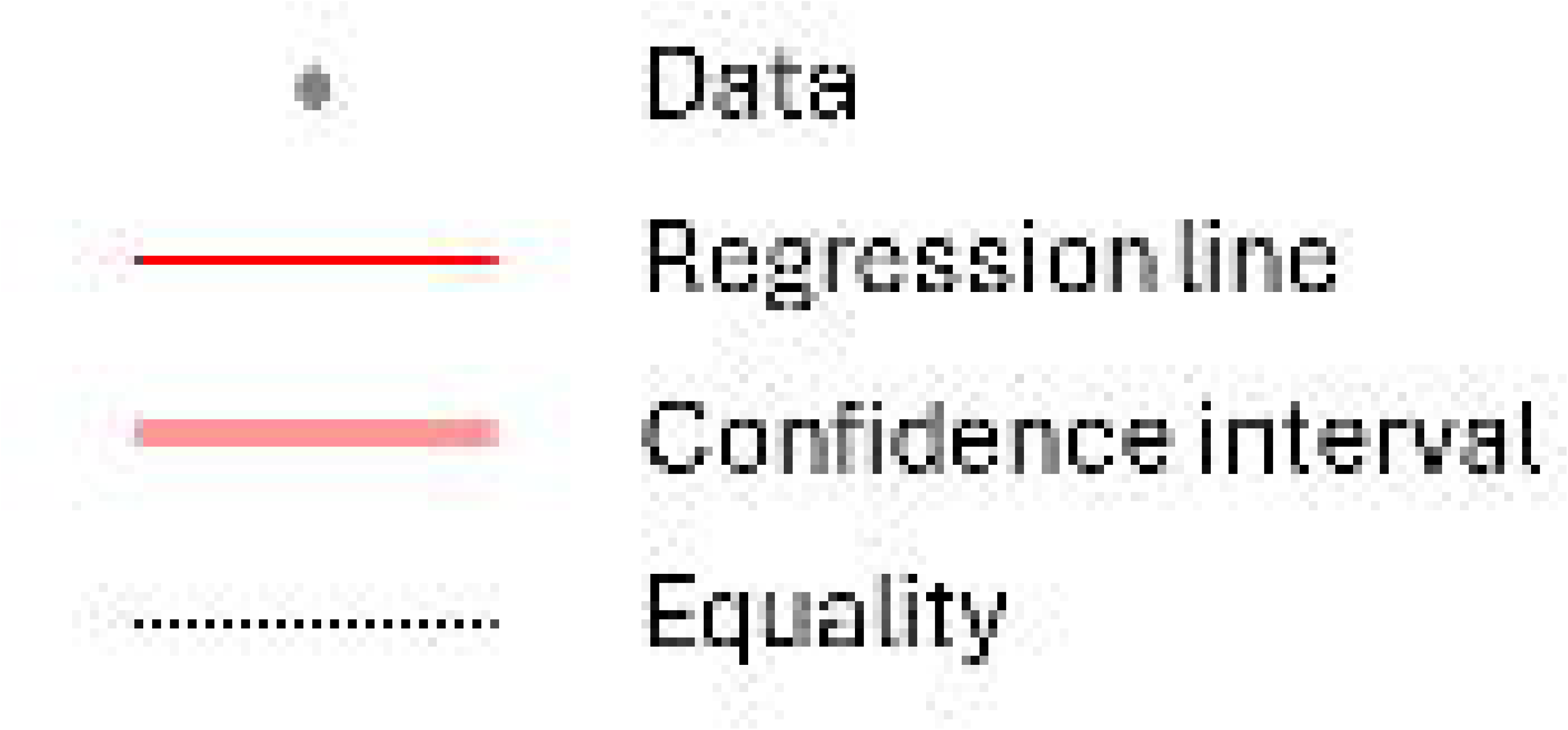
Comparison between the proportion of the population in the estimated catchment area and the proportion of actual respiratory admissions for the period 2017-2024 (hospitalisation) and 2019-2024 (emergency). Each point represents a department of residence, age group and year, with the value being the proportion of the total in that age group and year which are in that department. The emergency visit analysis was conducted only on departments in the ARA region. The Pearson correlation coefficient is shown on the top left, with the relationship and 95% confidence interval shown in the red line and the pink region around this showing the 95% confidence interval. The dashed line indicates where the two proportions are equal.

We also compared RSV incidence in hospitalised infants between the respiratory cohort at HCL and a national analysis (Figure 8), which demonstrated good agreement in terms of scale.

**Figure 8.**
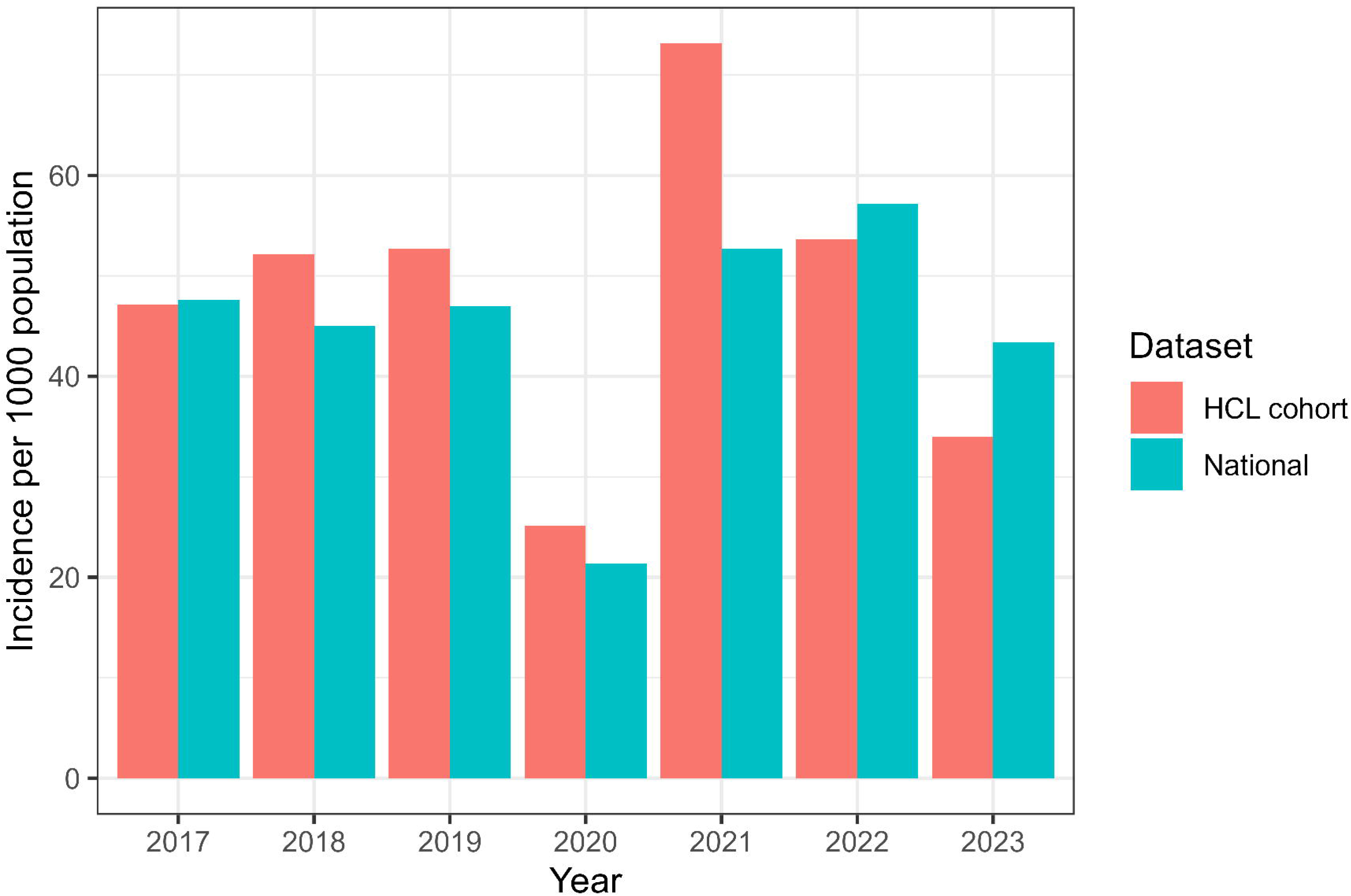
RSV incidence in hospitalised infants <1 year in the HCL respiratory cohort, using the catchment population estimated in this study as a denominator, compared with national incidence estimates (23).

## Discussion

This analysis leveraged extensive national data on numbers of hospitalisations in order to estimate time- and age-specific catchment populations for major hospital groups in Metropolitan France using a proportional flow framework. The total estimated catchment population across all hospital groups was stable over time and closely aligned with national population estimates, supporting the internal consistency of the approach. We estimated catchment populations for individual hospital groups that varied substantially by group, the largest being AP-HP, which had substantial numbers of hospitalisation visits from every department in Metropolitan France. We demonstrated the geographic proximity of the catchment area for the largest hospitals. For French hospitals and hospital groups, these estimates can serve as denominators for hospital-level data, providing a robust basis for comparing hospital-based disease incidence and for strengthening surveillance systems that rely on hospital case counts. We also estimated the distribution of catchment populations by the type of hospital, which may serve as a rule of thumb to provide catchment population estimates in other contexts where the data to estimate proportional flow are not available.

We explored the framework in detail using the HCL group as a case study. We calculated age-specific catchment populations, and found that these were similar over time suggesting that hospital preference patterns are relatively robust to short-term fluctuations in healthcare utilisation. We demonstrated that our catchment population estimates were considerably lower than the two possible census areas which could be used as denominators, the Rhone department and Metropolitan Lyon, suggesting that incidence estimates using those denominators underestimate the overall incidence. The discrepancy between the overall catchment population when estimated in an age-disaggregated analysis and when using aggregated ages demonstrates that aggregation may obscure important variation in hospital utilisation by age group. This could result in either under- or over-estimates of denominators. Given that many surveillance systems report age-stratified incidence or severity estimates, age-specific denominator estimation should be preferred where feasible.

The choice of geographic resolution also materially influenced denominator estimates. In principle, the use of smaller geographic areas is appropriate where possible because Bailey’s method assumes that hospital admission rates and utilisation patterns within each geographic area are constant (2). Greater spatial disaggregation may therefore better reflect true variation in hospital preference patterns and provide a more accurate representation of the population served. Furthermore, when catchment estimation is used not only for incidence calculation but also to characterise the health needs of the population served by a hospital, finer geographic resolution may offer additional insight (26). However, the finer spatial resolution increased the requirement to mask data. We therefore selected the department as the geographic level of analysis as a default resolution, as it provided a pragmatic balance between spatial specificity and minimal data exclusion in age-stratified analyses.

Catchment population sizes for the HCL differed markedly by diagnostic category and visit type, although whether this reflects the attractiveness of the HCL for these types of conditions, or the extent to which people will travel for treatment, requires further analysis. Emergency department visits were associated with more localised catchment populations compared with complete hospital admissions. These findings demonstrate that hospital denominators are context-specific and that a single catchment definition may not be appropriate for all surveillance or burden estimation objectives.

The strong correlation between the geographic distribution of the estimated catchment population, and the distribution of actual admissions in a respiratory cohort, supports the validity of this method of estimation even when the catchment populations were estimated from data on all causes of hospitalisation. This validation provides empirical support for the use of proportional flow–derived denominators in hospital-based infectious disease surveillance. We also validated the estimated catchment size by comparing RSV incidence nationally and at the HCL. This is based on the assumptions that incidence at HCL reflects national incidence, and that the denominator population for national incidence statistics is reliable as it need not contend with variation in choice of hospital within France. The similar magnitude of the resulting incidence measures supports the appropriateness of our denominator estimates.

Defining hospital geographic catchment areas remains methodologically challenging, particularly when fine granularity of geography or patient characteristics are required (7). In France, prior studies have used geographic distance to estimate healthcare accessibility (27,28), and to estimate catchment population to inform epidemic forecasting models (29). To our knowledge, application of a proportion flow approach using comprehensive administrative hospital data has not previously been described.

In lower resource settings, well-defined registers of residential addresses may be lacking, presenting challenges to hospitals to identify patient provenance in detail (30). Often, catchment population estimation in these settings has taken advantage of wider sources of data. A study in Uganda estimated catchment populations by including administrative areas according to three different measures: crow-fly distance from hospital, travel distance from hospital or comparison of disease incidence with overall hospital rates (31). Others have combined geographic data and hospital admissions data to estimate catchment populations in Kenya, but in the absence of information from all competing hospitals, a Bayesian approach was used to predict contributions of different areas based on travel time to the hospital (32). In contrast, the present framework benefits from complete national coverage of hospital admissions, allowing direct estimation of observed patient flows.

Our work is subject to certain limitations. We make the assumption that all individuals in any population in Metropolitan France will seek treatment if they require hospitalisation. If this is not the case, this would cause us to overestimate the relevant catchment area, and therefore to underestimate disease burdens in such under-served populations.

We validated the catchment population estimates using a cohort with a specific set of patients, those with acute respiratory illness. It is therefore possible that the correlation we demonstrated would not hold for a different type of cohort, particularly one which affected a more specific subset of the population (33).

A further limitation of our approach, which relies on prior data on patient flows to specific health facilities, is that it cannot account for a changing environment such as the opening of a new hospital, or the expansion or contraction of a current one. Gravity models, which take into account geographic distance and population size, provide a way to predict the effect of such a change (7), but require detailed data to calibrate them to the healthcare environment.

In conclusion, the availability of comprehensive hospital utilisation data combined with a proportional flow framework enables estimation of hospital catchment population denominators applicable to wide range of surveillance and burden estimation objectives.

Future work on this dataset could integrate travel time and other geographic accessibility measures with patient flows. This would allow validation of such assumptions as the gravity model, and allow prediction of how the catchment populations would evolve given a change in the healthcare environment. Furthermore the age, disease prevalence and socio-economic status in the source population as well as the type of hospital visited are also key sources of heterogeneity which merit further investigation.

## Data Availability

The datasets from ATIH and Urg'ARA which were analysed during the current study are available from Github at https://github.com/georgeshirreff/ZoneCap.
The respiratory cohort data are available from the Department of Health Data at the HCL, but restrictions apply to the availability of these data, which were used under license for the current study, and so are not publicly available. Data are however available from the authors upon reasonable request and with permission of the Department of Health Data.

## Declarations

### Ethics approval and consent to participate

The research on data from the HCL received authorisation from the Commission Nationale de l’Informatique et des Libertés (CNIL) number 923273 on 06/11/2023. Requirement for individual patient consent was waived in favour of an opt-out system whereby the details of the inclusion criteria for patients were published at least one month before their use and patients could therefore request to have their data excluded.

Data from ATIH and Urg’ARA were collected for routine purposes, and were provided for this study in aggregate, and masked at low numbers, thereby minimising the risk of any information being identifiable to specific individuals.

### Consent for publication

Not applicable

### Availability of data and materials

The datasets from ATIH and Urg’ARA which were analysed during the current study are available from Github at https://github.com/georgeshirreff/ZoneCap.

The respiratory cohort data are available from the Department of Health Data at the HCL, but restrictions apply to the availability of these data, which were used under license for the current study, and so are not publicly available. Data are however available from the authors upon reasonable request and with permission of the Department of Health Data.

### Authors’ contributions

G.S. conceived the principal analysis, conducted the analysis, drafted and finalised the manuscript. C.C. developed the methodology, implemented the creation of the respiratory cohort and edited the manuscript. J.-S.C. and P.V. provided scientific input on the creation of the respiratory cohort, on the methodology and edited the manuscript. C.D. implemented the creation of the respiratory cohort and edited the manuscript. A.R. acquired and managed the emergency visit data. K.T. developed the methodology and edited the manuscript. M.N. developed the methodology, oversaw the creation of the respiratory cohort and edited the manuscript. All authors read and approved the final manuscript.

## Acknowledgements

Not applicable

## Abbreviations

ATIH: Agence Technique de l’Information sur l’Hospitalisation
AP-HP: Assistance Publique–Hôpitaux de Paris
AP-HM: Assistance Publique–Hôpitaux de Marseille
ARA: Auvergne-Rhône-Alpes
CH: Centre Hospitalier
CHRU: Regional University Hospital Centre
CHU: University Hospital Centre
FINESS: Fichier National des Établissements Sanitaires et Sociaux
GCS: Groupement de coopération sanitaire
GH: Groupe Hospitalier
HCL: Hospices Civils de Lyon
ICD-10: International Classification of Disease version 10
INSEE: Institut National de la Statistique et des Etudes Economiques
PCR: Polymerase Chain Reaction
PMSI: Programme de Médicalisation des Systèmes d’Information
SA: Société Anonyme

