## Supplementary Information for "Estimation of hospital catchment populations using data on patient hospital use in France"

### Enrolment diagnostic codes of the retrospective cohort on respiratory infections at HCL

A patient receiving any of the following exact ICD-10 codes could meet the criteria for enrolment:

J09, J13, J14, J22, J44.0, A48.1, B01.2, B05.2, B25.0, B37.1, B38.0, B39.0, B40.0, B41.0, B42.0, B44.0, B45.0, B46.0, B58.3, B59, U071.0, U071.1.

In addition, a patient could be enrolled if they received a code beginning with the following characters:

J10, J11, J12, J15, J16, J17, J18, J20, J21, J85, J86, P23.

Supplementary Table 1. Average annual number of hospitalisations and size of estimated catchment population from and catchment area within Metropolitan France for the 200 largest hospital groups, by number of hospitalisations.

| FINESS – Hospital group | Annual hospitali-sations | Estimated catchment population | Estimated catchment area |
| --- | --- | --- | --- |
| 750712184 - AP-HP | 525 515 | 3 999 251 | 4 999 |
| 690781810 - HOSPICES CIVILS DE LYON | 154 342 | 1 097 187 | 4 407 |
| 130786049 - AP-HM | 112 620 | 709 650 | 3 703 |
| 310781406 - CHU TOULOUSE | 112 118 | 799 420 | 7 409 |
| 330781196 - CHU DE BORDEAUX | 100 988 | 704 499 | 6 036 |
| 590780193 - CHRU DE LILLE | 92 191 | 567 661 | 1 614 |
| 670780055 - CHU DE STRASBOURG | 84 053 | 577 094 | 2 592 |
| 440000289 - CHU DE NANTES | 74 331 | 588 517 | 3 320 |
| 340780477 - CHU MONTPELLIER | 71 919 | 489 841 | 3 946 |
| 540023264 - CHRU DE NANCY | 69 514 | 444 182 | 4 582 |
| 760780239 - CHU ROUEN | 69 386 | 461 390 | 2 684 |
| 380780080 - CHU GRENOBLE ALPES | 67 414 | 486 231 | 3 408 |
| 350005179 - CHRU DE RENNES | 64 430 | 488 244 | 3 442 |
| 370000481 - CHU DE TOURS | 62 550 | 428 783 | 5 369 |
| 570005165 - CHR METZ THIONVILLE | 60 954 | 398 711 | 2 617 |
| 630780989 - CHU CLERMONT-FERRAND | 56 657 | 388 365 | 5 841 |
| 860014208 - CHU DE POITIERS | 56 261 | 390 555 | 6 193 |
| 800000044 - CHU D'AMIENS | 54 299 | 339 118 | 3 352 |
| 490000031 - CHRU ANGERS | 52 758 | 384 486 | 3 595 |
| 140000100 - CHU CAEN | 52 657 | 348 354 | 3 237 |
| 770021145 - GRAND HÔPITAL DE L'EST FRANCILIEN | 52 333 | 410 555 | 1 679 |
| 210780581 - CHU DIJON BOURGOGNE | 51 658 | 337 185 | 6 094 |
| 060785011 - CHU DE NICE | 51 102 | 329 729 | 1 495 |
| 290000017 - CHU BREST | 51 017 | 324 457 | 2 481 |
| 420784878 - CHU SAINT ETIENNE | 50 369 | 315 355 | 2 519 |
| 250000015 - CHU BESANCON | 47 507 | 339 228 | 4 240 |
| 870000015 - CHU LIMOGES | 45 902 | 288 786 | 5 442 |
| 590782215 - CH VALENCIENNES | 45 851 | 283 795 | 659 |
| 720000025 - CH LE MANS | 45 838 | 292 419 | 3 334 |
| 450000088 - CHU D'ORLEANS | 45 210 | 318 885 | 3 436 |
| 510000029 - CHR DE REIMS | 43 683 | 287 438 | 4 466 |
| 300780038 - CHU NIMES | 42 887 | 291 884 | 2 305 |
| 740781133 - CH ANNECY-GENEVOIS | 42 460 | 317 290 | 2 036 |
| 850000019 - CHD VENDEE | 42 126 | 290 159 | 2 761 |
| 730000015 - CH METROPOLE SAVOIE | 40 789 | 270 340 | 3 499 |
| 750000523 - GROUPEMENT HOSPITALIER PARIS SAINT-JOSEPH | 40 399 | 312 636 | 251 |
| 830100616 - CHIC TOULON | 39 242 | 239 647 | 1 318 |
| 680020336 - GRPE HOSP REGION MULHOUSE ET SUD ALSACE | 37 920 | 270 276 | 1 261 |
| 130785652 - HÔPITAL SAINT JOSEPH | 37 753 | 236 945 | 1 047 |
| 840006597 - CH HENRI DUFFAUT AVIGNON | 36 766 | 241 327 | 1 472 |
| 910002773 - CH SUD-FRANCILIEN | 36 402 | 273 893 | 487 |
| 560005746 - GROUPEMENT HOSPITALIER BRETAGNE SUD | 36 081 | 239 301 | 2 053 |
| 900000365 - HÔPITAL NORD FRANCHE COMTE | 34 996 | 258 117 | 2 240 |
| 680000973 - CH DE COLMAR | 34 468 | 244 335 | 1 150 |
| 220000020 - CH DE SAINT BRIEUC PAIMPOL TREGUIER | 33 835 | 207 293 | 2 348 |
| 950110080 - HÔPITAL NOVO | 33 665 | 259 438 | 433 |
| 560023210 - CH BRETAGNE ATLANTIQUE VANNES | 33 225 | 222 828 | 1 950 |
| 660780180 - CH PERPIGNAN | 32 326 | 223 062 | 1 928 |
| 760780726 - GROUPEMENT HOSPITALIER DU HAVRE | 31 989 | 212 664 | 1 126 |
| 290020700 - CHIC DE CORNOUAILLE QUIMPER | 30 348 | 193 928 | 1 418 |
| 590782421 - CH DE ROUBAIX | 30 017 | 186 161 | 413 |
| 170024194 - GROUPEMENT HOSPITALIER DE LA ROCHELLE-RE-AUNIS | 29 545 | 199 031 | 2 065 |
| 780110078 - CH DE VERSAILLES | 28 633 | 229 960 | 386 |
| 130041916 - CH DU PAYS D'AIX CHI AIX PERTUIS | 28 460 | 179 113 | 1 078 |
| 260000021 - CH VALENCE | 28 182 | 183 218 | 2 443 |
| 640780417 - CH COTE BASQUE | 28 008 | 192 798 | 2 584 |
| 920000650 - HÔPITAL FOCH | 27 572 | 216 104 | 214 |
| 620100685 - CH DE LENS | 27 060 | 163 769 | 732 |
| 330781253 - CH DE LIBOURNE | 26 603 | 184 684 | 1 625 |
| 690782222 - CH NORD OUEST VILLEFRANCHE | 26 176 | 189 808 | 649 |
| 440000057 - CH ST-NAZAIRE | 26 020 | 210 980 | 1 033 |
| 790000012 - CH DE NIORT | 25 767 | 170 869 | 2 659 |
| 280000134 - CH CHARTRES | 25 696 | 165 018 | 2 233 |
| 640781290 - CH DE PAU | 25 431 | 174 756 | 2 068 |
| 770170017 - CH MARNE LA VALLEE | 25 350 | 173 201 | 702 |
| 950110015 - CH VICTOR DUPOUY ARGENTEUIL | 25 092 | 196 212 | 207 |
| 780001236 - CHIC DE POISSY ST-GERMAIN | 25 056 | 201 959 | 346 |
| 590781415 - CH DE DUNKERQUE | 25 033 | 155 361 | 356 |
| 270023724 - CH EURE SEINE | 24 772 | 167 227 | 1 631 |
| 910110055 - GROUPEMENT HOSPITALIER NORD ESSONNE | 24 477 | 184 598 | 256 |
| 940110018 - CHIC DE CRETEIL | 24 409 | 184 370 | 106 |
| 600101984 - GROUPEMENT HOSPITALIER PUBLIC DU SUD DE L'OISE | 24 151 | 164 859 | 1 110 |
| 950013870 - GHEM EAUBONNE MONTMORENCY SIMONE VEIL | 24 039 | 187 092 | 190 |
| 080011174 - CHIC NORD ARDENNES | 23 994 | 155 007 | 3 064 |
| 100000017 - CH DE TROYES | 23 716 | 161 702 | 3 118 |
| 710780958 - CH WILLIAM MOREY CHALON SUR SAONE | 23 516 | 131 809 | 2 073 |
| 020000063 - CH DE SAINT QUENTIN | 23 396 | 137 564 | 1 810 |
| 440041580 - L'HÔPITAL PRIVÉ DU CONFLUENT | 22 877 | 180 425 | 1 011 |
| 350000022 - GROUPEMENT HOSPITALIER RANCE EMERAUDE | 22 675 | 170 151 | 1 211 |
| 620103440 - CH DE BOULOGNE | 22 582 | 136 664 | 624 |
| 590783239 - CH DE DOUAI | 22 506 | 138 733 | 341 |
| 770700185 - CH DE MEAUX | 22 481 | 152 675 | 671 |
| 950110049 - CH DE GONESSE | 22 444 | 173 381 | 190 |
| 310780259 - SA CLINIQUE PASTEUR | 22 404 | 156 421 | 1 766 |
| 930110051 - CH DE ST DENIS | 22 355 | 167 213 | 45 |
| 940110042 - CHIC DE VILLENEUVE ST GEORGES | 22 184 | 167 252 | 156 |
| 510024979 - POLYCLINIQUE DE BEZANNES | 22 098 | 148 197 | 2 304 |
| 620100057 - CH D'ARRAS | 21 610 | 130 774 | 605 |
| 710780263 - CH LES CHANAUX MACON | 21 598 | 129 832 | 1 709 |
| 340780055 - CH BEZIERS | 21 586 | 148 215 | 837 |
| 240000117 - CH DE PERIGUEUX | 21 470 | 136 433 | 2 989 |
| 930110069 - CH ROBERT BALLANGER | 21 380 | 159 880 | 104 |
| 770110054 - GRPE HOSPITALIER DU SUD ILE DE FRANCE | 21 281 | 166 673 | 679 |
| 600100721 - CHIC COMPIEGNE-NOYON | 21 267 | 144 411 | 1 039 |
| 330780479 - POLYCLINIQUE BX-NORD AQUITAINE | 21 204 | 150 339 | 1 032 |
| 500000013 - CH DU COTENTIN | 21 132 | 131 958 | 1 587 |
| 890000037 - CH AUXERRE | 20 979 | 120 683 | 2 721 |
| 160000451 - CH ANGOULEME | 20 618 | 142 922 | 2 416 |
| 080000615 - CH CHARLEVILLE MEZIERES | 20 304 | 112 504 | 2 164 |
| 190000042 - CH DUBOIS BRIVE | 20 302 | 122 682 | 3 037 |
| 350000121 - CH PRIVÉ ST-GREGOIRE | 20 302 | 153 169 | 1 092 |
| 490000676 - CH CHOLET | 20 279 | 146 054 | 1 383 |
| 600100713 - CH DE BEAUVAIS | 20 197 | 137 745 | 967 |
| 740790258 - CH ALPES LEMAN | 20 193 | 151 800 | 893 |
| 010780054 - CH BOURG EN BRESSE | 20 179 | 145 138 | 1 330 |
| 670780337 - CH DE HAGUENAU | 20 113 | 138 617 | 595 |
| 110780061 - CH CARCASSONNE | 19 701 | 117 734 | 1 973 |
| 690805361 - CH ST JOSEPH ST LUC | 19 470 | 140 535 | 338 |
| 620100651 - CH BETHUNE-BEUVRY | 19 374 | 117 464 | 528 |
| 770021152 - CH SUD SEINE ET MARNE | 19 299 | 150 060 | 695 |
| 760024042 - CHIC ELBEUF LOUVIERS VAL DE REUIL | 19 161 | 128 421 | 1 010 |
| 440033819 - SANTÉ ATLANTIQUE | 19 085 | 151 904 | 846 |
| 700004591 - GROUPEMENT HOSPITALIER DE HAUTE SAONE | 19 064 | 119 437 | 2 692 |
| 930110036 - CH ANDRE GREGOIRE | 18 741 | 140 323 | 36 |
| 180000028 - CH J. COEUR BOURGES | 18 649 | 119 584 | 2 918 |
| 590797353 - GCS GHICL HÔPITAL SAINT VINCENT | 18 628 | 115 833 | 269 |
| 920300043 - HÔPITAL PRIVÉ D ANTONY | 18 430 | 140 609 | 110 |
| 310780283 - NOUVELLE CLINIQUE DE L'UNION | 18 429 | 130 652 | 1 080 |
| 060780988 - CH DE CANNES | 18 316 | 118 569 | 468 |
| 580780039 - CH DE L'AGGLOMÉRATION DE NEVERS | 18 266 | 100 198 | 3 206 |
| 930021480 - GHI LE RAINCY-MONTFERMEIL | 18 264 | 136 424 | 154 |
| 590781902 - CH DE TOURCOING | 18 199 | 112 536 | 253 |
| 420011413 - HÔPITAL PRIVÉ DE LA LOIRE | 17 958 | 112 030 | 900 |
| 750150104 - INSTITUT MUTUALISTE MONTSOURIS | 17 825 | 135 221 | 224 |
| 380780049 - CH PIERRE OUDOT | 17 699 | 128 138 | 788 |
| 360000053 - CH DE CHATEAUROUX | 17 637 | 110 005 | 3 432 |
| 170780175 - GH SAINTES - SAINT JEAN D'ANGELY | 17 450 | 118 348 | 1 258 |
| 400011177 - CHIC MONT DE MARSAN ET PAYS DES SOURCES | 17 403 | 116 392 | 2 660 |
| 400780193 - CH DAX - COTE D'ARGENT | 17 336 | 117 306 | 2 508 |
| 310781000 - CLINIQUE DES CEDRES | 17 265 | 121 441 | 1 304 |
| 410000087 - CH DE BLOIS | 17 043 | 114 851 | 2 183 |
| 120780044 - CH RODEZ | 17 041 | 113 247 | 3 543 |
| 430000018 - CH LE PUY | 16 945 | 100 592 | 2 257 |
| 870000288 - CLINIQUE FRANÇOIS CHENIEUX | 16 917 | 106 225 | 1 994 |
| 620101337 - CH DE CALAIS | 16 902 | 102 556 | 449 |
| 750006728 - GROUPEMENT HOSPITALIER DIACONESSES-CROIX SAINT-SIMON | 16 823 | 116 809 | 76 |
| 030780118 - CH VICHY | 16 612 | 95 655 | 2 004 |
| 670018068 - CLINIQUE RHENA GCS | 16 470 | 116 514 | 511 |
| 260000047 - GROUPEMENT HOSPITALIER PORTES DE PROVENCE | 16 429 | 106 652 | 1 409 |
| 650783160 - CH TARBES LOURDES | 16 330 | 105 129 | 2 018 |
| 280000183 - CH VICTOR JOUSSELIN | 16 287 | 106 152 | 1 326 |
| 370007569 - PÔLE SANTÉ LÉONARD DE VINCI | 16 274 | 112 103 | 1 232 |
| 050002948 - CHICAS GAP-SISTERON | 16 204 | 96 248 | 3 628 |
| 490014909 - CLINIQUE DE L'ANJOU | 16 184 | 118 786 | 1 066 |
| 420780033 - CH ROANNE | 16 077 | 100 847 | 670 |
| 590780268 - POLYCLINIQUE DU BOIS | 16 069 | 99 238 | 250 |
| 620101360 - CH REGION DE SAINT-OMER - SITE D'HELFAUT | 15 981 | 96 682 | 421 |
| 060780954 - CH ANTIBES-JUAN LES PINS | 15 973 | 103 744 | 408 |
| 500000054 - HÔPITAUX DU SUD MANCHE | 15 942 | 100 322 | 1 183 |
| 380012658 - GROUPEMENT HOSPITALIER MUTUALISTE DE GRENOBLE | 15 889 | 114 333 | 729 |
| 130043664 - HÔPITAL EUROPEEN DESBIEF AMBROISE PARE | 15 777 | 98 796 | 350 |
| 310026927 - CLINIQUE CAPIO LA CROIX DU SUD | 15 714 | 118 226 | 902 |
| 810000380 - CHIC CASTRES-MAZAMET | 15 623 | 99 420 | 1 371 |
| 830100566 - CHIC FREJUS | 15 603 | 95 758 | 519 |
| 940000664 - INSTITUT GUSTAVE ROUSSY | 15 502 | 114 014 | 469 |
| 530000371 - CH LAVAL | 15 322 | 101 894 | 1 717 |
| 690041132 - MEDIPOLE HÔPITAL MUTUALISTE | 15 254 | 116 121 | 269 |
| 880007059 - CHIC EMILE DURKHEIM EPINAL | 15 160 | 85 918 | 1 406 |
| 500000112 - CH SAINT-LO | 15 110 | 94 843 | 1 113 |
| 800000028 - CH D'ABBEVILLE | 15 082 | 93 390 | 945 |
| 020000261 - CH DE SOISSONS | 15 064 | 87 962 | 1 201 |
| 340015502 - CLINIQUE LE MILLENAIRE | 15 054 | 103 214 | 712 |
| 780110011 - CH FRANCOIS QUESNAY MANTES LA JOLIE | 15 046 | 120 459 | 251 |
| 130789316 - CH DE MARTIGUES | 15 036 | 94 713 | 249 |
| 470016171 - CH AGEN-NERAC | 14 959 | 92 313 | 1 523 |
| 760780023 - CH DIEPPE | 14 924 | 99 170 | 508 |
| 380781435 - CH VIENNE | 14 822 | 105 754 | 609 |
| 060780715 - CLINIQUE SAINT GEORGE | 14 803 | 94 998 | 425 |
| 060780897 - CH DE GRASSE | 14 798 | 95 447 | 395 |
| 030780100 - CH DE MONTLUCON NERIS-LES-BAINS | 14 747 | 84 118 | 1 985 |
| 890970569 - CH SENS | 14 522 | 85 380 | 1 783 |
| 210012670 - HÔPITAL PRIVÉ DIJON BOURGOGNE | 14 513 | 97 998 | 1 744 |
| 140000035 - CH LISIEUX | 14 364 | 95 910 | 841 |
| 130001647 - INSTITUT PAOLI - CALMETTES | 14 349 | 90 006 | 704 |
| 590780284 - GCS GHICL HÔPITAL ST PHILIBERT | 14 338 | 88 634 | 219 |
| 720017748 - CENTRE MÉDICO-CHIRURGICAL DU MANS | 14 236 | 89 975 | 1 050 |
| 590780227 - GROUPEMENT HOSPITALIER SECLIN CARVIN | 14 224 | 87 398 | 256 |
| 300780046 - CH ALES | 14 202 | 96 560 | 878 |
| 390780146 - CH JURA SUD | 14 143 | 86 856 | 1 632 |
| 590781803 - CH SAMBRE-AVESNOIS | 14 103 | 87 593 | 203 |
| 560014748 - CH DU CENTRE BRETAGNE | 13 933 | 89 380 | 877 |
| 610780082 - CHIC ALENCON MAMERS | 13 887 | 79 770 | 1 441 |
| 540000478 - NOUVELLE CLINIQUE LOUIS PASTEUR | 13 858 | 86 255 | 1 086 |
| 450000104 - CH AGGLOMERATION MONTARGOISE | 13 815 | 97 956 | 978 |
| 940000649 - HÔPITAL SAINT-CAMILLE | 13 800 | 104 679 | 85 |
| 570000646 - HÔPITAL CLINIQUE CLAUDE BERNARD | 13 732 | 88 503 | 592 |
| 2A0000014 - CH GAL D'AJACCIO | 13 701 | 89 959 | 2 124 |
| 140017237 - HÔPITAL PRIVÉ ST MARTIN | 13 701 | 89 411 | 866 |
| 2B0000020 - CH BASTIA | 13 692 | 97 446 | 2 441 |
| 740790381 - CHIC LES HÔPITAUX DU LEMAN | 13 666 | 102 616 | 588 |
| 130782634 - CH DE SALON | 13 655 | 85 878 | 227 |
| 170780225 - CH ROCHEFORT | 13 587 | 91 339 | 944 |
| 640018206 - CLINIQUE BELHARRA | 13 461 | 92 157 | 1 395 |
| 370000093 - SAS NOUVELLE CLINIQUE DE TOURS + | 13 448 | 94 004 | 1 184 |
| 820000016 - CH MONTAUBAN | 13 415 | 87 751 | 1 242 |
| 630781839 - CLINIQUE CHATAIGNERAIE - BEAUMONT | 13 383 | 91 597 | 1 426 |
| 570026252 - HÔPITAL ROBERT SCHUMAN | 13 358 | 88 384 | 585 |
| 590781605 - CH DE CAMBRAI | 13 335 | 82 437 | 213 |
| 030780092 - CH MOULINS YZEURE | 13 329 | 74 780 | 1 700 |
| 110780137 - CH NARBONNE | 13 289 | 80 040 | 1 278 |

Supplementary Table 2. Average annual number of hospitalisations and size of estimated catchment population from and catchment area within Metropolitan France for the 100 largest individual hospitals, by number of hospitalisations.

| Individual hospital | FINESS – Hospital group | Annual hospitalis-ations | Estimated catchment population | Estimated catchment area |
| --- | --- | --- | --- | --- |
| 760000158 - HÔPITAL CHARLES NICOLLE CHU ROUEN | 760780239 - CHU ROUEN | 62 090 | 413 428 | 2 423 |
| 330781360 - GROUPEMENT HOSPITALIER PELLEGRIN - CHU | 330781196 - CHU DE BORDEAUX | 60 198 | 421 436 | 3 467 |
| 750100125 - GHU APHP SUN SITE PITIE SALPETRIERE | 750712184 - AP-HP | 60 054 | 452 608 | 906 |
| 380000067 - HÔPITAL NORD - CHU38 | 380780080 - CHU GRENOBLE ALPES | 57 257 | 410 566 | 2 927 |
| 440000271 - CHU DE NANTES SITE HOTEL DIEU HME | 440000289 - CHU DE NANTES | 54 841 | 435 007 | 2 418 |
| 490000049 - CHU ANGERS SITE LARREY | 490000031 - CHRU ANGERS | 52 758 | 384 486 | 3 595 |
| 140000209 - CHU CAEN | 140000100 - CHU CAEN | 51 690 | 341 936 | 3 185 |
| 860000223 - CHU LA MILETRIE | 860014208 - CHU DE POITIERS | 50 396 | 347 875 | 5 512 |
| 800006124 - CHU AMIENS SUD | 800000044 - CHU D'AMIENS | 49 854 | 312 182 | 3 079 |
| 210987558 - HÔPITAL LE BOCAGE CHRU DIJON | 210780581 - CHU DIJON BOURGOGNE | 49 706 | 323 890 | 5 874 |
| 420785354 - HÔPITAL NORD - CHU42 | 420784878 - CHU SAINT ETIENNE | 47 880 | 299 585 | 2 394 |
| 250006954 - CHU JEAN MINJOZ BESANCON | 250000015 - CHU BESANCON | 46 678 | 333 200 | 4 166 |
| 590000618 - CH VALENCIENNES | 590782215 - CH VALENCIENNES | 45 851 | 283 795 | 659 |
| 720000033 - CH DU MANS | 720000025 - CH LE MANS | 45 565 | 290 553 | 3 312 |
| 450002613 - CHRU ORLEANS - HÔPITAL DE LA SOURCE | 450000088 - CHU D'ORLEANS | 44 114 | 311 024 | 3 315 |
| 750100166 - GHU APHP CUP SITE COCHIN PORT ROYAL | 750712184 - AP-HP | 42 961 | 327 607 | 442 |
| 350000741 - CHRU RENNES SITE PONTCHAILLOU | 350005179 - CHRU DE RENNES | 42 798 | 321 905 | 2 318 |
| 300782117 - CHU NIMES CAREMEAU | 300780038 - CHU NIMES | 42 544 | 289 448 | 2 286 |
| 130783293 - APHM HÔPITAL LA TIMONE | 130786049 - AP-HM | 42 464 | 267 798 | 1 469 |
| 670783273 - HÔPITAL DE HAUTEPIERRE | 670780055 - CHU DE STRASBOURG | 40 700 | 280 095 | 1 264 |
| 750000523 - GROUPEMENT HOSPITALIER PARIS SAINT-JOSEPH | 750000523 - GROUPEMENT HOSPITALIER PARIS SAINT-JOSEPH | 40 399 | 312 636 | 251 |
| 310783048 - HÔPITAL PURPAN CHU TOULOUSE | 310781406 - CHU TOULOUSE | 38 227 | 274 203 | 2 313 |
| 570026682 - HÔPITAL DE MERCY - CHR METZ THIONVILLE | 570005165 - CHR METZ THIONVILLE | 38 084 | 248 988 | 1 688 |
| 540002698 - CHRU NANCY - HÔPITAUX DE BRABOIS | 540023264 - CHRU DE NANCY | 37 898 | 240 039 | 2 675 |
| 940100043 - GHU APHP UPS SITE KREMLIN BICETRE APHP | 750712184 - AP-HP | 37 776 | 284 475 | 351 |
| 130785652 - HÔPITAL SAINT JOSEPH | 130785652 - HÔPITAL SAINT JOSEPH | 37 753 | 236 945 | 1 047 |
| 850000142 - CHD SITE LA ROCHE SUR YON | 850000019 - CHD VENDEE | 36 829 | 253 715 | 2 421 |
| 910020254 - CH SUD FRANCILIEN SITE JEAN JAURES | 910002773 - CH SUD-FRANCILIEN | 36 402 | 273 893 | 487 |
| 840001861 - CH D'AVIGNON HENRI DUFFAUT | 840006597 - CH HENRI DUFFAUT AVIGNON | 36 261 | 238 015 | 1 446 |
| 310783055 - HÔPITAL DE RANGUEIL CHU TOULOUSE | 310781406 - CHU TOULOUSE | 36 151 | 255 481 | 2 700 |
| 730000031 - CHMS - SITE CHAMBERY MCO | 730000015 - CH METROPOLE SAVOIE | 36 145 | 239 697 | 3 103 |
| 690784152 - HÔPITAL CROIX-ROUSSE | 690781810 - HOSPICES CIVILS DE LYON | 35 715 | 256 573 | 897 |
| 590796975 - HÔPITAL SALENGRO CHU LILLE | 590780193 - CHRU DE LILLE | 35 638 | 219 570 | 619 |
| 870000064 - CHU DUPUYTREN LIMOGES | 870000015 - CHU LIMOGES | 35 327 | 222 016 | 4 237 |
| 130780521 - APHM HÔPITAL NORD | 130786049 - AP-HM | 35 199 | 221 588 | 1 047 |
| 830000345 - CHITS CH SAINTE MUSSE | 830100616 - CHIC TOULON | 34 963 | 213 581 | 1 173 |
| 670000025 - HÔPITAL CIVIL / NOUVEL HÔPITAL CIVIL | 670780055 - CHU DE STRASBOURG | 34 638 | 237 487 | 1 068 |
| 630000404 - HÔPITAL GABRIEL MONTPIED - CHU63 | 630780989 - CHU CLERMONT-FERRAND | 34 485 | 235 854 | 3 587 |
| 740000237 - CH ANNECY-GENEVOIS SITE ANNECY | 740781133 - CH ANNECY-GENEVOIS | 34 474 | 258 335 | 1 600 |
| 750100232 - GHU APHP NUP SITE BICHAT C BERNARD | 750712184 - AP-HP | 34 027 | 259 602 | 206 |
| 900003039 - HNFC SITE TREVENANS | 900000365 - HÔPITAL NORD FRANCHE COMTE | 33 213 | 249 232 | 2 162 |
| 940100027 - GHU APHP HM SITE HENRI MONDOR | 750712184 - AP-HP | 33 058 | 249 686 | 341 |
| 220000012 - CH YVES LE FOLL | 220000020 - CH DE SAINT BRIEUC PAIMPOL TREGUIER | 32 950 | 201 483 | 2 284 |
| 690784137 - HÔPITAL LYON SUD | 690781810 - HOSPICES CIVILS DE LYON | 32 621 | 231 272 | 892 |
| 660000084 - CH PERPIGNAN | 660780180 - CH PERPIGNAN | 32 326 | 223 062 | 1 928 |
| 680000684 - HÔPITAL LOUIS PASTEUR | 680000973 - CH DE COLMAR | 32 260 | 229 966 | 1 083 |
| 950000364 - HÔPITAL NOVO SITE PONTOISE | 950110080 - HÔPITAL NOVO | 31 913 | 245 359 | 394 |
| 560000135 - GHBS- HÔPITAL DU SCORFF | 560005746 - GROUPEMENT HOSPITALIER BRETAGNE SUD | 31 697 | 210 269 | 1 828 |
| 760805770 - HÔPITAL JACQUES MONOD CH LE HAVRE | 760780726 - GROUPEMENT HOSPITALIER DU HAVRE | 31 638 | 210 311 | 1 114 |
| 690783154 - HÔPITAL EDOUARD HERRIOT | 690781810 - HOSPICES CIVILS DE LYON | 31 416 | 223 956 | 839 |
| 680004546 - HÔPITAL EMILE MULLER | 680020336 - GRPE HOSP REGION MULHOUSE ET SUD ALSACE | 31 363 | 225 591 | 1 054 |
| 560000127 - CHBA SITE DE VANNES | 560023210 - CH BRETAGNE ATLANTIQUE VANNES | 31 007 | 208 075 | 1 820 |
| 290004324 - CHRU BREST SITE HÔPITAL CAVALE BLANCHE | 290000017 - CHU BREST | 30 201 | 192 247 | 1 457 |
| 170000087 - HÔPITAL SAINT-LOUIS - LA ROCHELLE | 170024194 - GROUPEMENT HOSPITALIER DE LA ROCHELLE-RE-AUNIS | 29 529 | 198 933 | 2 064 |
| 750803447 - GHU APHP CUP SITE G POMPIDOU HEGP | 750712184 - AP-HP | 29 329 | 224 313 | 306 |
| 290000025 - CHIC QUIMPER | 290020700 - CHIC DE CORNOUAILLE QUIMPER | 29 108 | 186 058 | 1 360 |
| 780800256 - CH DE VERSAILLES SITE ANDRE MIGNOT | 780110078 - CH DE VERSAILLES | 28 633 | 229 955 | 386 |
| 590801106 - HÔPITAL VICTOR PROVO | 590782421 - CH DE ROUBAIX | 28 227 | 175 841 | 390 |
| 260000013 - CH DE VALENCE | 260000021 - CH VALENCE | 28 182 | 183 218 | 2 443 |
| 370000861 - CHRU BRETONNEAU - TOURS | 370000481 - CHU DE TOURS | 28 180 | 193 614 | 2 395 |
| 060785003 - CHU DE NICE HÔPITAL PASTEUR | 060785011 - CHU DE NICE | 28 150 | 183 060 | 836 |
| 640000162 - CH COTE BASQUE | 640780417 - CH COTE BASQUE | 28 008 | 192 798 | 2 584 |
| 920000650 - HÔPITAL FOCH | 920000650 - HÔPITAL FOCH | 27 572 | 216 104 | 214 |
| 750100208 - GHU CUP SITE NECKER ENFANTS MALADES | 750712184 - AP-HP | 27 554 | 209 201 | 402 |
| 750100042 - GHU APHP NUP SITE LARIBOISIERE | 750712184 - AP-HP | 27 537 | 210 544 | 166 |
| 370004467 - CHRU TROUSSEAU - CHAMBRAY | 370000481 - CHU DE TOURS | 27 328 | 186 866 | 2 355 |
| 330783648 - HÔPITAL HAUT-LEVEQUE - CHU | 330781196 - CHU DE BORDEAUX | 27 263 | 188 955 | 1 841 |
| 620000257 - CH LENS | 620100685 - CH DE LENS | 27 060 | 163 769 | 732 |
| 690000575 - CH NORD OUEST - VILLEFRANCHE | 690782222 - CH NORD OUEST VILLEFRANCHE | 26 176 | 189 808 | 649 |
| 330000605 - CH DE LIBOURNE -R.BOULIN | 330781253 - CH DE LIBOURNE | 26 152 | 181 550 | 1 589 |
| 440000016 - CH DE SAINT NAZAIRE | 440000057 - CH ST-NAZAIRE | 25 972 | 210 553 | 1 031 |
| 790000087 - CH DE NIORT | 790000012 - CH DE NIORT | 25 767 | 170 869 | 2 659 |
| 280504267 - CH CHARTRES LOUIS PASTEUR-LE COUDRAY | 280000134 - CH CHARTRES | 25 696 | 165 018 | 2 233 |
| 640000600 - CH DE PAU | 640781290 - CH DE PAU | 25 431 | 174 756 | 2 068 |
| 770019032 - CH DE MARNE LA VALLEE SITE JOSSIGNY | 770170017 - CH MARNE LA VALLEE | 25 350 | 173 201 | 702 |
| 950000307 - CH VICTOR DUPOUY | 950110015 - CH VICTOR DUPOUY ARGENTEUIL | 25 092 | 196 212 | 207 |
| 130000409 - CHIC SITE D'AIX EN PROVENCE | 130041916 - CH DU PAYS D'AIX CHI AIX PERTUIS | 24 906 | 156 232 | 901 |
| 750100273 - GHU APHP SUN SITE TENON | 750712184 - AP-HP | 24 492 | 186 682 | 128 |
| 940000573 - CHIC DE CRETEIL | 940110018 - CHIC DE CRETEIL | 24 409 | 184 370 | 106 |
| 590000337 - CH DUNKERQUE | 590781415 - CH DE DUNKERQUE | 24 369 | 151 617 | 347 |
| 310016977 - HÔPITAUX MERE ET ENFANTS CHU TOULOUSE | 310781406 - CHU TOULOUSE | 24 308 | 176 737 | 1 312 |
| 950000323 - GHEM SIMONE VEIL SITE EAUBONNE | 950013870 - GHEM EAUBONNE MONTMORENCY SIMONE VEIL | 24 039 | 187 092 | 190 |
| 770019032 - GHEF MARNE LA VALLEE SITE JOSSIGNY | 770021145 - GRAND HÔPITAL DE L'EST FRANCILIEN | 23 546 | 185 243 | 727 |
| 710978263 - CH WILLIAM MOREY CHALON SUR SAONE | 710780958 - CH WILLIAM MOREY CHALON SUR SAONE | 23 516 | 131 809 | 2 073 |
| 020000162 - CH SAINT-QUENTIN | 020000063 - CH DE SAINT QUENTIN | 23 396 | 137 564 | 1 810 |
| 690007539 - HÔPITAL FEMME MERE ENFANT | 690781810 - HOSPICES CIVILS DE LYON | 23 300 | 166 959 | 603 |
| 100000090 - CH DE TROYES | 100000017 - CH DE TROYES | 23 260 | 158 590 | 3 057 |
| 440041580 - L'HÔPITAL PRIVÉ DU CONFLUENT | 440041580 - L'HÔPITAL PRIVÉ DU CONFLUENT | 22 877 | 180 425 | 1 011 |
| 750100091 - GHU APHP SUN SITE ST ANTOINE | 750712184 - AP-HP | 22 718 | 172 702 | 180 |
| 620000653 - CH BOULOGNE-SUR-MER | 620103440 - CH DE BOULOGNE | 22 582 | 136 664 | 624 |
| 340785161 - HÔPITAL LAPEYRONIE CHU MONTPELLIER | 340780477 - CHU MONTPELLIER | 22 581 | 153 907 | 1 161 |
| 780000311 - CHIC POISSY ST GERMAIN SITE DE POISSY | 780001236 - CHIC DE POISSY ST-GERMAIN | 22 572 | 183 298 | 315 |
| 340796663 - HÔPITAL ARNAUD DE VILLENEUVE CHU MPT | 340780477 - CHU MONTPELLIER | 22 522 | 153 790 | 1 210 |
| 590001004 - CH DOUAI DECHY | 590783239 - CH DE DOUAI | 22 506 | 138 733 | 341 |
| 770000446 - CH DE MEAUX SITE SAINT FARON | 770700185 - CH DE MEAUX | 22 481 | 152 675 | 671 |
| 950000331 - CH GENERAL DE GONESSE | 950110049 - CH DE GONESSE | 22 444 | 173 381 | 190 |
| 310780259 - SA CLINIQUE PASTEUR | 310780259 - SA CLINIQUE PASTEUR | 22 404 | 156 421 | 1 766 |
| 940000599 - CHIC LUCIE ET RAYMOND AUBRAC | 940110042 - CHIC DE VILLENEUVE ST GEORGES | 22 184 | 167 252 | 156 |
| 510024979 - POLYCLINIQUE DE BEZANNES | 510024979 - POLYCLINIQUE DE BEZANNES | 22 098 | 148 197 | 2 304 |
| 930000328 - CH GENERAL DELAFONTAINE | 930110051 - CH DE ST DENIS | 22 075 | 164 962 | 44 |

Supplementary Table 3. Average annual number of hospitalisations and size of estimated catchment population from and catchment area within overseas regions for the 50 largest hospital groups, by number of hospitalisations.

| FINESS – Hospital group | Annual  hospitalisations | Estimated catchment population | Estimated catchment area |
| --- | --- | --- | --- |
| 970408589 - CHU DE LA REUNION | 66 430 | 473 999 | 1 343 |
| 970211207 - CHU DE MARTINIQUE | 37 167 | 295 227 | 1 690 |
| 970306809 - GCS PREFIGURATEUR DU CHU DE GUYANE | 29 141 | 222 377 | 63 546 |
| 980500003 - CH DE MAYOTTE | 27 949 | 263 558 | 339 |
| 970100228 - CHU DE LA GUADELOUPE | 22 392 | 169 702 | 779 |
| 970302022 - CH DE CAYENNE | 17 571 | 124 860 | 36 991 |
| 970421038 - CH OUEST REUNION | 17 346 | 121 828 | 350 |
| 970462107 - CLINIQUE SAINTE CLOTILDE | 16 455 | 117 139 | 330 |
| 970403606 - GH EST REUNION | 11 529 | 81 712 | 234 |
| 970302121 - CH FRANK JOLY | 11 047 | 78 675 | 23 208 |
| 970107249 - CLINIQUE LES EAUX CLAIRES | 8 107 | 60 679 | 266 |
| 970100178 - CH DE LA BASSE TERRE | 7 968 | 61 230 | 263 |
| 970300265 - CENTRE MÉDICO-CHIRURGICAL DE KOUROU | 5 661 | 39 685 | 12 324 |
| 970202313 - CLINIQUE SAINT PAUL | 5 076 | 40 451 | 138 |
| 970100012 - POLYCLINIQUE DE LA GUADELOUPE | 4 514 | 34 866 | 148 |
| 970305629 - CHIC DE KOUROU | 4 497 | 32 063 | 9 373 |
| 970462073 - CLINIQUE DURIEUX | 4 255 | 29 837 | 86 |
| 970462081 - CLINIQUE LES ORCHIDEES | 4 210 | 30 161 | 86 |
| 970100186 - CH LOUIS CONSTANT FLEMING | 3 383 | 144 | 47 |
| 750712184 - AP-HP | 3 122 | 23 141 | 1 483 |
| 970462024 - CLINIQUE JEANNE D'ARC | 2 541 | 17 477 | 51 |
| 970202321 - SA CLINIQUE SAINTE MARIE | 2 184 | 16 494 | 48 |
| 970100020 - CENTRE MEDICO-SOCIAL | 1 846 | 14 287 | 61 |
| 970404844 - CLINIQUE SAINT-VINCENT | 912 | 6 435 | 18 |
| 970302055 - HÔPITAL PRIVÉ SAINT-GABRIEL | 887 | 6 308 | 1 888 |
| 970202156 - HÔPITAL DU MARIN | 878 | 6 977 | 21 |
| 970102596 - CLINIQUE DE CHOISY | 855 | 6 593 | 35 |
| 940000664 - INSTITUT GUSTAVE ROUSSY | 710 | 5 377 | 259 |
| 970100202 - CH SAINTE-MARIE | 685 | 5 318 | 23 |
| 970100160 - HL IRÉNÉE DE BRUYN | 573 | 0 | 11 |
| 970404109 - INSTITUT ROBERT DEBRE | 483 | 3 371 | 10 |
| 970103099 - CLINIQUE LES NOUVELLES EAUX-MARINES | 473 | 3 654 | 16 |
| 970202164 - HÔPITAL DE SAINT-ESPRIT | 461 | 3 658 | 11 |
| 970100137 - POLYCLINIQUE SAINT-CHRISTOPHE | 388 | 3 004 | 13 |
| 970100244 - CH DE CAPESTERRE-BELLE-EAU, EX H.L. | 224 | 1 697 | 7 |
| 970305843 - GUYANE SANTÉ HIBISCUS | 147 | 1 051 | 303 |
| 130786049 - AP-HM | 144 | 1 142 | 35 |
| 750000549 - FONDATION OPHTALMOLOGIQUE ROTHSCHILD | 143 | 1 088 | 82 |
| 970100285 - CH M.SELBONNE | 143 | 1 077 | 5 |
| 920000684 - CENTRE CHIRURGICAL MARIE LANNELONGUE | 141 | 1 119 | 49 |
| 690781810 - HOSPICES CIVILS DE LYON | 137 | 1 046 | 58 |
| 750000523 - GROUPEMENT HOSPITALIER PARIS SAINT-JOSEPH | 126 | 946 | 42 |
| 330781196 - CHU DE BORDEAUX | 117 | 869 | 44 |
| 750150104 - INSTITUT MUTUALISTE MONTSOURIS | 88 | 670 | 36 |
| 750160012 - CLCC INSTITUT CURIE | 87 | 659 | 41 |
| 310781406 - CHU TOULOUSE | 86 | 642 | 53 |
| 950630012 - HÔPITAL D ENFANTS MARGENCY | 86 | 652 | 85 |
| 970107454 - A.U.D.R.A. | 78 | 437 | 2 |
| 750300154 - CLINIQUE TURIN | 74 | 540 | 83 |
| 690000880 - CENTRE LEON BERARD | 72 | 511 | 152 |
